# Resting-state EEG network markers differentiate people with epilepsy and functional seizures

**DOI:** 10.64898/2026.04.14.26350505

**Authors:** Peter Kissack, Wessel Woldman, Rachel Sparks, Joel S. Winston, Franz Brunnhuber, Naima Ciulini, Allan H. Young, Irene Faiman, Paul Shotbolt

## Abstract

**Background:** Distinguishing epilepsy from functional/dissociative seizures (FDS) is an ongoing diagnostic challenge. Using a well-controlled clinical EEG dataset, we run the first diagnostic accuracy study assessing the potential of resting-state EEG network markers to directly discriminate between the two conditions at the time when a diagnosis is suspected and prior to treatment initiation.

**Methods:** The dataset, previously examined in a published study, includes 148 age- and sex-matched individuals with suspected seizure disorder, later diagnosed with non-lesional epilepsy (n=75) or FDS (n=73). Functional network measures in the 6-9 Hz range were extracted from normal-looking, eyes-closed resting-state EEG data acquired while participants were medication-free. Machine learning was implemented to assess their predictive potential; different model configurations were tested to identify the most promising approach for translational implementations.

**Results:** EEG-derived network measures discriminate between conditions at levels significantly above chance (maximum balanced accuracy: 65.8%; SD: 2.9). Their sensitivity to epilepsy (78.4%; SD: 5.8)) is higher than their sensitivity to FDS (53.1%; SD: 4.0). Multiple nonlinear models succeed on the classification problem, but model choice remains a determinant of overall accuracy.

**Conclusion:** We establish evidence for the clinical validity of selected network-based markers to discriminate between a diagnosis of non-lesional epilepsy and FDS prior to treatment initiation, highlighting the measures’ potential to support post-test probability estimation in the clinic. These measures are more specific to epilepsy than FDS and should not be interpreted as markers of a positive diagnosis of FDS.

## INTRODUCTION

Paroxysmal episodes involving loss of consciousness require careful investigation, as these may be a symptom of epilepsy or semiologically similar disorders such as functional/dissociative seizures (FDS). Misdiagnosis and long diagnostic delays are common in these populations, and associated with significant risks,[1–3] compromising timely access to appropriate treatment and symptom control, with detrimental effects on morbidity, psychosocial well-being[4,5] and a substantial economic burden.[6]

While meticulous history taking, understanding or witnessing seizure semiology and clinical investigations such as electroencephalogram (EEG) and magnetic resonance imaging (MRI) can inform decision-making, diagnosis often remains a clinical judgement.[7,8] Consistently, the diagnostic sensitivity of a single routine EEG after a first unprovoked epileptic seizure is estimated at 17% in adults[9], and is likely even lower in FDS, where capture of a typical event during a short EEG assessment is infrequent.

Identifying biomarkers that can aid the diagnosis of seizure disorders is an ongoing effort.[10] One line of work focuses on extrapolating quantitative information from routine EEGs to identify markers from visually normal interictal activity that would otherwise be clinically inconclusive.[11] The challenge is identifying features reliably correlating with diagnostic traits, rather than transitory clinical states, medication effects, comorbidities, or demographic information.[11,12] Previous work demonstrated that single-channel EEG features show limited diagnostic validity in distinguishing epilepsy from FDS when these confounders are controlled for within a diagnostic accuracy design.[12]

However, despite findings often conflicting at the individual measure-level, evidence suggests alterations in functional connectivity and network measures in people with epilepsy, indicating discriminative patterns might emerge in the multivariate space.[11] This aligns with theories highlighting the role of synchronisation in seizure generation;[13] a network approach may better capture network-level dynamics potentially affecting synchronisability. Recent studies comparing people with epilepsy to siblings, healthy controls,[14–19] or epilepsy mimics (including FDS)[20,21] provide further evidence for network measures. Increasingly larger samples and stronger designs capture the variability expected in clinical practice[20] especially when care is taken in quantifying confounders’ effects.[15–17,20]

However, the direct differentiation between non-lesional epilepsy and FDS remains underexplored,[11] offering an opportunity for tackling a diagnostic problem of direct clinical relevance, where even modest improvements to the diagnostic yield of routine EEGs[9] could be valuable. Building on recent progress and earlier findings for this dataset,[12] we evaluate the potential of network measures to directly discriminate between the two seizure disorders using a well-controlled dataset. Demographic matching and confounder control at the design stage, including absence of CNS agents and major comorbidities, allow robust exploration of diagnostic markers and mechanistic understanding. We therefore emphasise identifying which modelling decisions hold greatest promise for future translational implementations.

Even with inconclusive clinical EEG findings, a diagnostic decision-support tool based on resting-state EEGs could contribute to post-test probability estimation, aid non-specialist settings in acknowledging diagnostic uncertainty, support treatment initiation decisions prior to diagnosis, and optimise patient referral to tertiary services, potentially reducing diagnostic delays. This work represents a further step in this direction.

## MATERIALS AND METHODS

### Participants and data preprocessing

We use the resting-state EEG dataset previously reported in Faiman *et al*.[12] This includes a quasi-consecutive, age- and sex-matched sample of adults (>18 years) presenting with a suspected seizure disorder, having normal CT/MRI, not taking any CNS agents at the time of EEG, and ultimately receiving an established specialist diagnosis of non-lesional epilepsy (n=75) or FDS (n=73). For epilepsy, this consisted of a clinical diagnosis in accordance with operational clinical definitions;[22] for FDS, a video-EEG supported diagnosis as per ILAE definitions[7], or in the absence of video-EEG confirmation, a strong clinical indication of FDS following review by both a specialist neurologist and neuropsychiatrist. Sampling was consecutive except for a small subset (n=24) identified to achieve age and sex matching (Supplementary Section 1).[12] An up-to-date review of hospital notes (July 2024) revealed no diagnostic revisions. As we aimed to identify diagnostic markers relevant to epilepsy and FDS, the sample had no history of other neurological disorders, moderate to severe neurodevelopmental disorders, or active and severe mental health disorders; detailed comorbidity criteria are in Supplementary Section 1.

Twenty-one channel clinical EEG data were acquired by the King’s College Hospital Department of Neurophysiology using NicoletOne EEG System v44 or Xltek EEG32U, as previously described.[12] A blinded trained band 6 clinical physiologist (N.C.) selected awake, eyes-closed, visually normal, interictal resting-state data (excluding any major artifacts, non-specific or epileptiform abnormalities). Preprocessing is detailed in Faiman *et al.*[12] This included Xltek downsampling to 256Hz and bandpass-filtering at 0.5 and 70Hz to match the NicoletOne acquisition settings; line noise removal, bad channel spherical interpolation and re-referencing to robust common average (PREP[23]); rank-adjusted Independent Component Analysis followed by IClabel for component rejection[24]; finally, segmentation into 20s non-overlapping segments (epochs) to maintain consistency with previous studies.[11]

De-identified data preprocessed at King’s College London were transferred to the University of Birmingham for further analysis, in compliance with the study’s ethical approval (London Queen Square Research Ethics Committee REC: 20/LO/0784, IRAS ID: 265164, 28/09/2021).

### Connectivity-based feature extraction

For each EEG epoch, a network is computed based on the pairwise functional connectivity between 21 EEG channels, bandpass filtered to 6-9Hz, selected as a frequency band of interest based on previous studies.[20,25–29] Although spanning mainly the classical theta band, the 6 – 9 Hz range is referred to as ‘low alpha’[30] to maintain consistency with previous work.[20,25–29] From this, we calculate six measures covering a range of network properties, selected based on previous evidence from the epilepsy literature[14–16,18–20,31–36]: mean clustering coefficient[37], mean functional connectivity, network efficiency[38], trophic incoherence[39], betweenness centrality[40], and critical coupling[32,33] (Supplementary Section 2). Since each individual has several EEG epochs, we take the epoch-wise mean of each measure, under the assumption that diagnostic markers should reflect relatively stable neurophysiological traits. This approach also avoids treating multiple epochs from the same individual as independent observations, which results in inflated performance estimates in models that do not account for repeated measures from the same subject.

### Classification

We assess whether the extracted features can discriminate between the diagnosis of epilepsy and FDS, and what model choices perform best.

Model performance was evaluated using nested cross-validation (Fig. 1), with repeated fold selection stratified by diagnosis, distinguishing between *model selection* – the minimum number of EEG epochs used for computing feature averages, what features are considered, and which type of model is being evaluated – and *hyperparameters* – internal parameter choices to be trained in the inner tier of cross-validation (Table 1; Supplementary Section 2.3). We report each model’s balanced accuracy, metric chosen to equally represent the effect of correctly or incorrectly classifying samples in the epilepsy or FDS classes.

**Figure 1.**
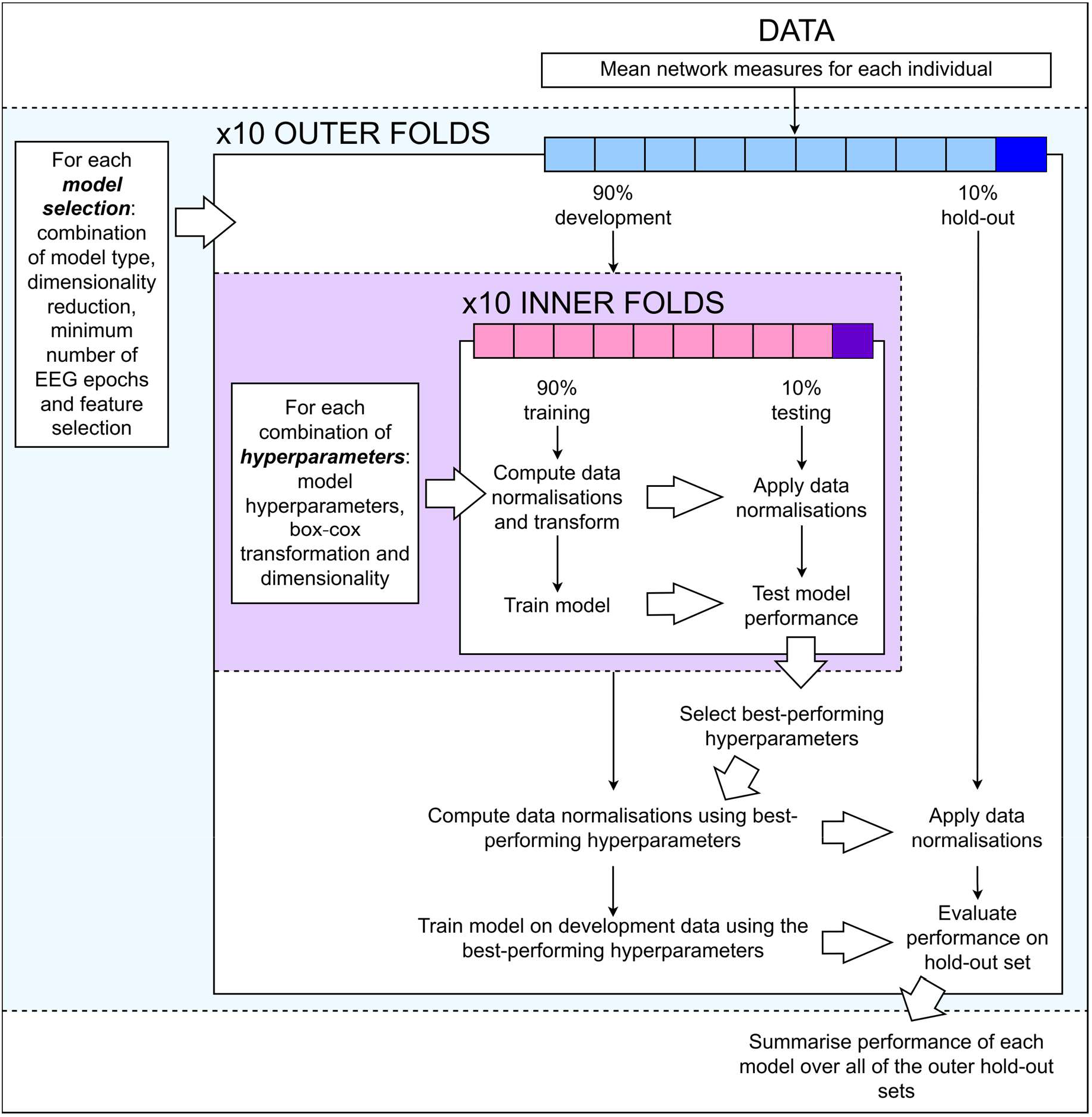
Outline of the nested cross-validation process. The outer tier, associated with the evaluation of the classification performance for a given model selection, is highlighted in blue. The inner tier, associated with the selection of hyperparameters within each development set, is highlighted in purple. Pre-specified search values for each hyperparameter are listed in Table 1. Narrow, downwards-pointing arrows represent the flow of data through the cross-validation procedure. Thick, white-filled arrows represent the transfer of parameter information between different subsets of the data.

**Table 1.** Summary of model selection and hyperparameters tested. In our nested cross-validation framework, model selection consists of the following considerations. A selection of model types is evaluated. Analyses are carried out with and without feature selection, and for three dimensionality reduction approaches, which may uncover underlying patterns which are latent in the raw features. Since each individual has a varying number of EEG epochs, we assess whether averaging from a greater number of epochs provides more stable feature estimates for improved classifier performance. We therefore run our analyses on both the full dataset, and the subset of individuals with at least four EEG epochs available. For each fold in the outer tier of cross-validation, we use the inner tier to search over a range of hyperparameter values associated to the specific model choices, including the implementation of Box-Cox transform, dimensionality (if dimensionality reduction is used in the current model configuration), and hyperparameters specific to individual model types.

| Model Selection and Hyperparameters |  |  |  |
| --- | --- | --- | --- |
| Model Configuration |  | Associated hyperparameter | Hyperparameter values |
| Minimum Number of EEG Epochs | 1 | N/A |  |
|  | 4 | N/A |  |
| Feature selection | No | N/A |  |
|  | Yes | N/A |  |
| N/A | | Box-Cox Transform | No transform, $\lambda$ to 0 d.p., 1 d.p. |
| Dimensionality Reduction | No Dimensionality Reduction | N/A |  |
|  | Principal Component Analysis | Dimensionality | 1,2,..., (#features - 1), No transform |
|  | Spectral Embedding | Dimensionality | 1,2,..., #features |
| Model Type | k-Nearest Neighbours | Number of neighbours | 3,5,7,11,15,20 |
|  |  | Weighting type | Uniform, Distance |
| | | $p$ | 1,2,3 |
|  | Logistic Regression | Penalty norm | None, L2 |
| | | $C$ | 0.1,1,10 |
| | Support Vector Machine (Linear Kernel) | $C$ | 0.1,1,10 |
| | Support Vector Machine (Radial Basis Function Kernel) | $C$ | 0.1,1,10 |
| | | $\gamma$ | 0.001,0.01,0.1,10,100,1000, "scale", "auto" |
| | Support Vector Machine (Sigmoid Kernel) | $C$ | 0.1,1,10 |
| | | $\gamma$ | 0.001,0.01,0.1,10,100,1000, "scale", "auto" |
| | | $r$ | -2.0,0.0,2.0 |
|  | Linear Discriminant Analysis | N/A |  |
|  | Multi-Layer Perceptron | Activation function | Identity, Logistic, tanh, ReLU |
|  |  | Hidden layer sizes | 25, 50, 100, 200, (25,25), (50,50), (100,100), (200,200), (100,50,25), (200,100,50), (200,100,50,25) |
|  |  | Optimisation solver | lbfgs, adam |
|  | Decision Tree | Maximum depth | None, 2,5,7,10,20 |
|  |  | Criterion | Gini, Log-loss, Entropy |
|  | Gaussian Naïve Bayes | N/A |  |
|  | Random Forest | Number of estimators | 100, 200, 500, 1000, 2000 |
|  |  | Maximum tree depth | None, 2,5,7,10,20 |
|  |  | Decision tree criterion | Gini, Log-loss, Entropy |
|  | AdaBoost | Number of estimators | 50, 100, 200, 500, 1000 |
|  |  | Maximum tree depth | None, 2,5,7,10,20 |
|  |  | Decision tree criterion | Gini, Log-loss, Entropy |
|  | RUSBoost | Number of estimators | 50, 100, 200, 500, 1000 |
|  |  | Maximum tree depth | None, 2,5,7,10,20 |
|  |  | Decision tree criterion | Gini, Log-loss, Entropy |
|  | XGBoost | Number of estimators | 50, 100, 200, 500, 1000 |
|  | Dummy Classifier | Strategy | Uniform, Most frequent |

We report the proportion of individuals for whom a classifier assigns the same classification across the ten repeats of fold selection, and the proportion for which the three best-performing classifiers achieve a consensus classification.

Within our differential diagnosis framework, we replace terminology of “sensitivity” and “specificity” with “sensitivity to the epilepsy class” and “sensitivity to the FDS class”, respectively. Conventionally, however, classification models require classes to be identified as positive and negative, respectively; we therefore code epilepsy as positive and FDS as negative.

### Baseline comparisons, subgroup performance, and alternative frequency bands

We run the nested cross-validation with classification labels randomly permuted (99 permutations chosen to balance robustness of the estimation and computational feasibility), and with demographic predictors only (age, sex and presence of comorbidities), to compare our network features’ performance against chance and non-EEG demographic and clinical data. For the best-performing model configuration, we report sensitivity and balanced accuracy for different subgroups (stratifying by sex, presence of comorbidities, presence of EEG abnormalities captured elsewhere in the recordings, epilepsy type, and whether a functional seizure was captured during the EEG appointment). Lastly, we run the same nested cross-validation framework using networks derived from classical frequency bands (delta, 1-3.5Hz, theta, 4-7.5Hz, alpha, 8-12.5Hz, beta, 13-30Hz).

### Additional analyses

We produce box-plots of raw features both for the full sample and the subset with at least four EEG epochs used in the average feature estimation. Mann-Whitney U-test is implemented to compare median raw feature values between groups. A binomial generalised linear model (GLM) with a logit link function is fitted to ensure potential confounders (age, sex, presence of comorbidities) are not predictive of diagnosis in the subset with four or more epochs. Chi-squared test is used to assess group differences in categorical variables (or Fisher’s exact test for categories containing fewer than five individuals); t-tests for continuous normally-distributed variables, and Mann-Whitney U-tests for continuous variables with other distributions. Watson-Williams test is implemented to compare means of circular data (time of day of EEG recording, in radians). References for statistical tests are in Supplementary Section 2.

## RESULTS

### Population characteristics

The characteristics of the full sample including 148 people with an eventually established diagnosis of epilepsy (n=75) or FDS (n=73) are summarised in Table 2, as previously reported.[12] One of the model configurations tested here involves only considering people with at least four EEG epochs; this subgroup consists of 102 people (epilepsy n=57, FDS n=45; Table 2).

**Table 2.**
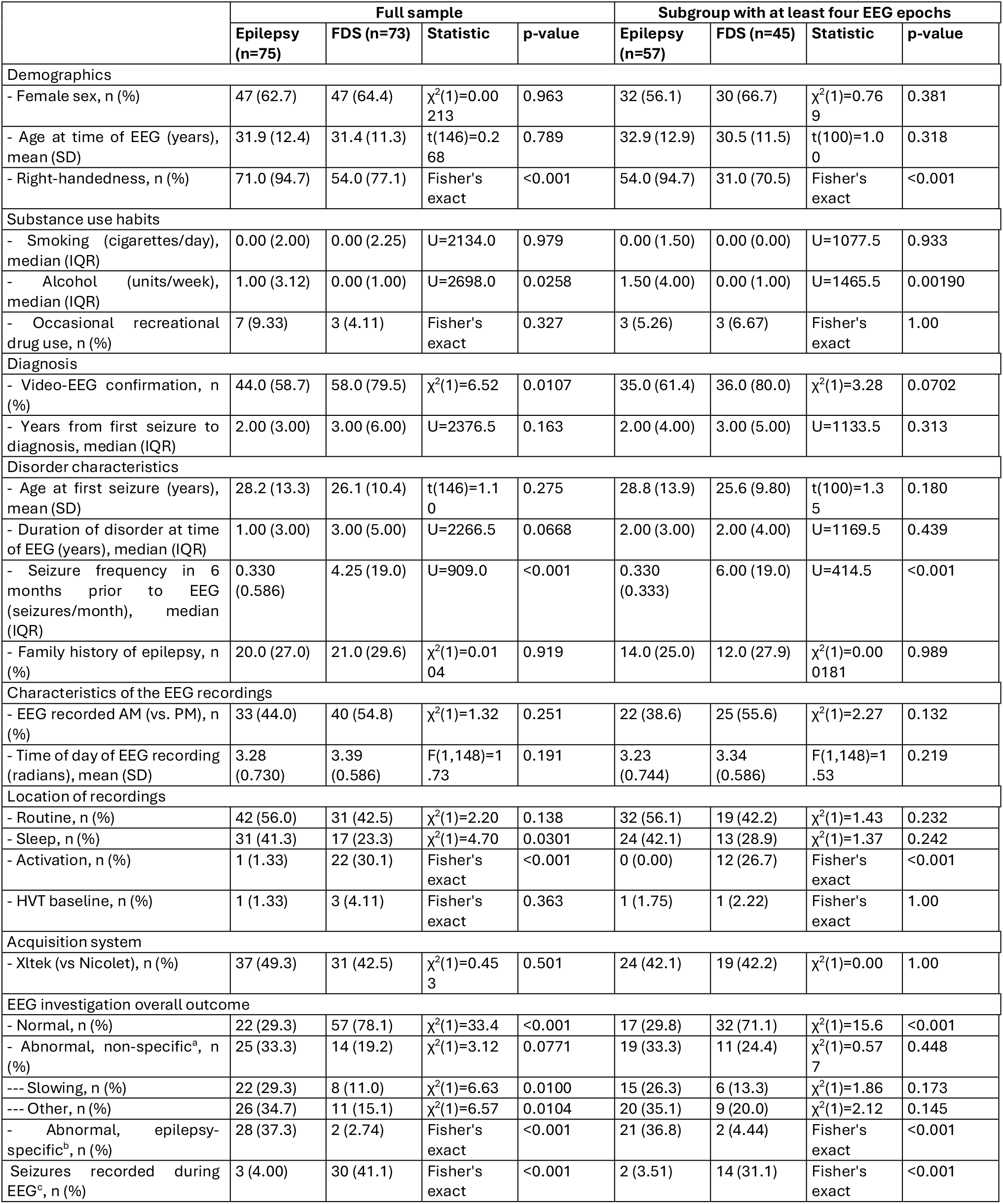
Demographic, clinical, and EEG-related characteristics. Demographic, clinical, and EEG-related characteristics are presented for the full sample and the subgroup with at least four EEG epochs used for average feature estimation, with statistical comparisons. ^a^Non-specific transient abnormalities refer to slowing or other non-specific transients (sharpened waves). ^b^Epilepsy-specific transient abnormalities refer to spikes, sharp waves, generalized spike and wave discharges, generalised polyspike-waves, generalized 3–4 Hz spikes. In the FDS group, specialist review concluded that these events were not attributable to epilepsy. ^c^ Seizures refer to epileptic seizures for the epilepsy group and functional seizures for the FDS group.

In both sample sets, most variables are balanced across diagnostic groups, except for a significantly higher number of left-handed or ambidextrous in the FDS group, higher monthly seizure frequency in the FDS group, and higher weekly alcohol consumption in the epilepsy group (Table 2). In both the epilepsy and FDS groups, approximately a third of the sample reported a family history of epilepsy. In the full epilepsy sample, 44% (n=33) have focal epilepsy, 25% (n=19) generalised epilepsy, and 31% (n=23) unknown or undefined; corresponding proportions in the subset with at least four EEG epochs are 49% (n=28), 28% (n=16), and 23% (n=13).

Groups do not differ significantly in the EEG acquisition system used nor in the time of the day when EEG recordings were taken. EEGs were performed between 2007-2021 for both groups. Analysed EEG segments contain no abnormalities based upon visual inspection.

The diagnosis is supported by video-EEG findings in 80% (subgroup: 80%) of people with FDS (typical event recorded in the absence of EEG abnormalities), and in 59% (subgroup: 61%) of those with epilepsy (epilepsy-specific abnormalities captured, not necessarily in the EEGs analysed here; Table 2).

### Classification

Balanced accuracy for distinguishing people with epilepsy from those with FDS in the full sample ranges from 51.7% to 62.9%, with variation dependent on model type (Supplementary Table S9).

Different model configurations are tested for their potential to improve classification performance (Table 1). The accuracy achieved is comparable (51.7-63.4%) when feature selection is implemented (Supplementary Tables S1, S3). Only considering the subset of people with at least four EEG epochs for average feature estimation (n=102) leads to a very marginal improvement in performance (51.2-65.8% without feature selection, Table 3; 51.0-64.1% with feature selection, Supplementary Tables S2, S6). Applying dimensionality reduction does not yield clear improvements (without feature selection, full sample, PCA: 51.4-61.5%, Supplementary Table S10; SE: 50.3-62.0%, Supplementary Table S11; subsample with ≥4 epochs, PCA: 51.1-65.0%, Supplementary Table S12; SE: 51.2-60.9%, Supplementary Table S13; with feature selection, full sample, PCA: 51.7-61.8%, Supplementary Table S4; SE: 50.0-60.0%, Supplementary Table S5; subsample with ≥4 epochs, PCA: 50.1-62.9%, Supplementary Table S7; SE: 48.7-58.5%, Supplementary Table S8). Lastly, model type is a key determinant of performance; the SVM with a Radial Basis Function kernel performs most consistently well across all configurations tested (57.4-65.8%); other nonlinear models (e.g., k-Nearest Neighbours, Random Forest, adaBoost, RUSboost, multi-layer perceptron) also perform well frequently (Table 3; Supplementary Tables S3-S13).

**Table 3.**
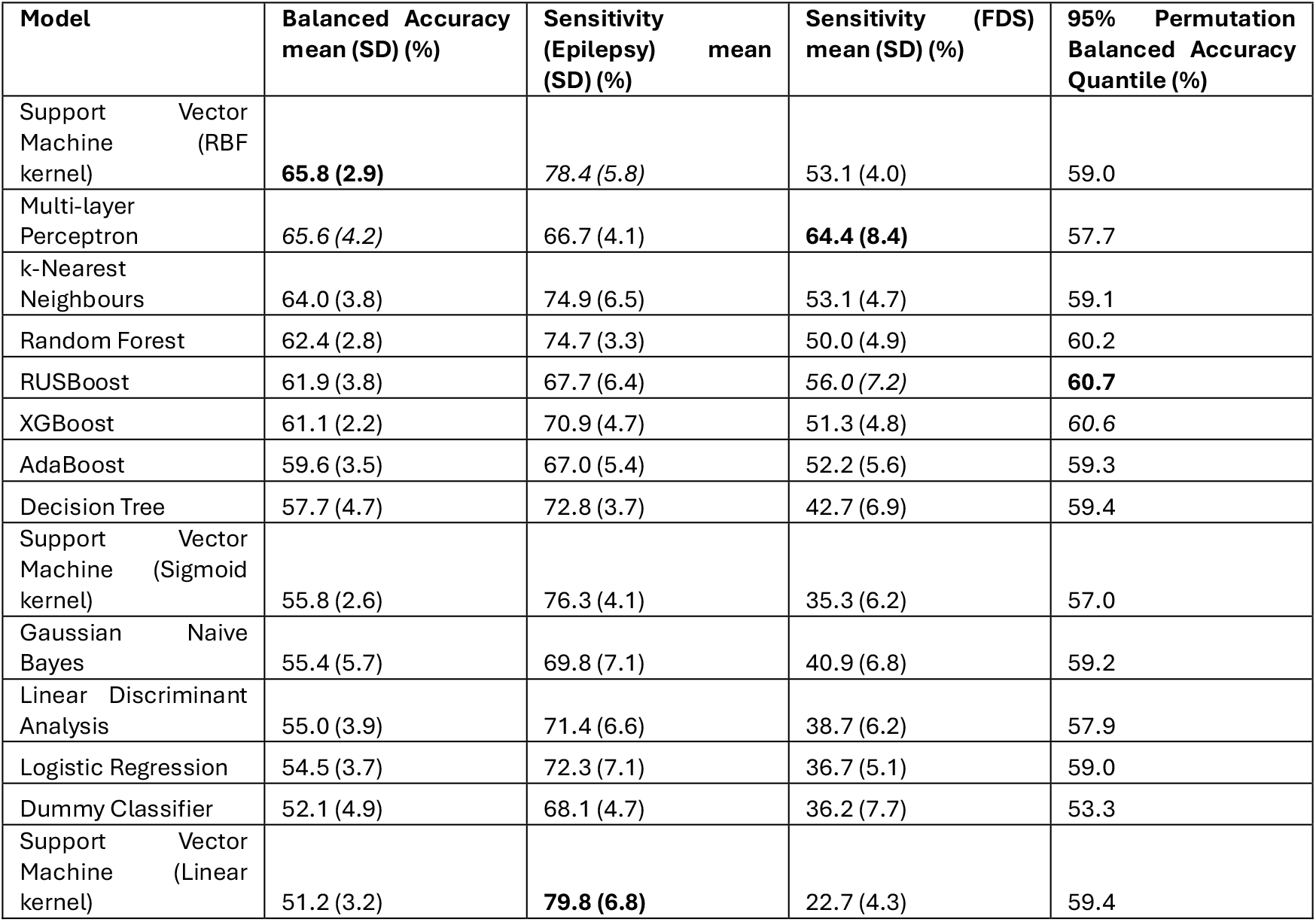
Results for the best-performing model configuration. The best-performing model configuration is based on using no feature selection or dimensionality reduction, and only considering individuals with at least four EEG epochs supporting average feature estimation (n = 102). For each of the ten repeats of fold selection, each individual is classified exactly once as part of one of the ten non-overlapping testing folds. Means and standard deviations are calculated for outcome measures across these ten repeats. The 95^th^ percentile (fifth highest) mean balanced accuracy obtained from repeating the same nested cross-validation process for each model type on 99 permutations of the true classification labels is reported in the final column. The best-performing value of each outcome measure is highlighted in bold; the second-best performing is in italic.

Consistently across all model configurations tested, the sensitivity for the epilepsy class is significantly higher than the sensitivity for the FDS class (Table 3).

Overall, the best-performing model configuration involves restricting analyses to people with at least four epochs for average feature estimation, applying no dimensionality reduction or feature selection, and using a SVM model with a Radial Basis Function (RBF) kernel; this achieves a final balanced accuracy of 65.8% with 78.4% sensitivity to the epilepsy class and 53.1% sensitivity to the FDS class (Table 3).

For the best configuration, classification consistency between the three best-performing models is explored. SVM-RBF provides the same classification across all ten repeats for 65.7% of people, Multi-Layer Perceptron for 40.2%, and k-Nearest Neighbours for 62.7%. These figures are independent of true diagnostic label. For 68.6% of people, the most frequently chosen classification is the same for the three best-performing models.

### Baseline comparisons, subgroup performance, and alternative frequency bands

For all cases where the balanced accuracy exceeds 60%, it also exceeds the 95^th^ percentile of permutation-based balanced accuracy (for SVM-RBF, 58.3%; Table 3). Using potentially confounding variables as predictors does not yield balanced accuracies higher than 59% for any model configuration.

Subgroup analyses indicate that classification performance is comparable across sexes, overall outcome of the EEG examination, epilepsy type, and across people with and without comorbidities (for the best performing configuration: Table 4; for full sample: Supplementary Table S14). Although classification performance appears higher for people with FDS in proximity to experiencing a functional seizure (60.0%) than those with no functional seizure captured during the EEG appointment (37.9%; Table 4), this difference is based on a small sample and is no longer evident when the full sample is considered (Supplementary Table S14).

**Table 4.** Subgroup analyses results. Balanced accuracy, sensitivity to epilepsy and sensitivity to FDS for characteristic subgroups of the data, using the best-performing model configuration.

| Subgroup | Size of epilepsy subgroup | Size of FDS subgroup | Balanced Accuracy (%) | Sensitivity (Epilepsy) (%) | Sensitivity (FDS) (%) |
| --- | --- | --- | --- | --- | --- |
| Sex, male | 25 | 15 | 68.8 | 81.6 | 56.0 |
| Sex, female | 32 | 30 | 63.8 | 75.9 | 51.7 |
| EEG normal | 17 | 32 | 66.3 | 84.7 | 47.8 |
| EEG abnormal, non-specific | 19 | 11 | 71.5 | 74.7 | 68.2 |
| EEG abnormal, epilepsy-specific | 21 | 2 | 65.8 | 76.7 | 55.0 |
| Comorbidity, absent | 43 | 26 | 69.3 | 76.3 | 62.3 |
| Comorbidity, present | 14 | 19 | 62.8 | 85.0 | 40.5 |
| Epilepsy type, focal | 28 | 0 | n/a | 82.1 | n/a |
| Epilepsy type, generalised | 16 | 0 | n/a | 75.0 | n/a |
| Epilepsy type, unknown | 11 | 0 | n/a | 80.0 | n/a |
| Functional seizure not captured during EEG appointment | 0 | 31 | n/a | n/a | 37.9 |
| Functional seizure captured during EEG appointment | 0 | 14 | n/a | n/a | 60.0 |

Detailed results for the nested cross-validation using networks derived from classical frequency bands are in Supplementary Section 4.3. Lower frequency bands showed higher balanced accuracy (six predictor models; delta: 48-58%; theta: 51-60%; Supplementary Tables S15-S16) than higher frequency bands (alpha: 40-52%; beta: 42-54%; Supplementary Tables S17-S18).

### Additional analyses

Mann-Whitney U-tests identify no significant group differences in the distributions of individual measures in either the full sample or subsample (Supplementary Table S19, Supplementary Fig. S1).

For the subsample, a GLM of diagnostic class against age, sex and the presence of comorbidities results in no statistically significant regression coefficients (Supplementary Table S20).

## DISCUSSION

For the first time, we assess whether network measures extracted from resting-state EEG can be implemented for the specific problem of directly differentiating between a diagnosis of epilepsy and one of functional/dissociative seizures, a common problem in clinical practice. We implement a machine-learning classification pipeline within a diagnostic accuracy design.

Leveraging clinical data from 148 non-lesional patients evaluated for seizure disorders prior to diagnosis, we show that network measures derived from interictal EEG can discriminate between conditions at levels significantly above chance (maximum balanced accuracy achieved: 65.8% in the 6-9Hz low alpha band).

Network measures have previously been used to characterise the epileptic brain compared to healthy controls[11,14–18,41] and to discriminate epilepsy from heterogeneous control groups including several differential diagnoses.[20,21] All previous studies included people on ASMs, and most included cohorts with longstanding, often drug-resistant epilepsy, whose findings may have limited applicability to individuals prior to diagnosis. For the first time, we establish evidence for the clinical validity of selected network measures and their potential to support diagnostic probability estimation at the time when non-lesional epilepsy or FDS is suspected, prior to treatment initiation.

We emphasise testing different model configurations to identify the most promising strategies for future translational work. First, we show that epoch-wise averaging, despite improving the temporal stability of network features, does not lead to a meaningful improvement in classification performance (62.9% to 65.8%). This, however, should be interpreted in the context of the accompanying moderate sample size reduction (n=148 to n=102), which may have limited the ability to detect greater improvements. Demographic and clinical variables remain balanced in this subgroup, showing no diagnostic predictive value in a control analysis. Therefore, while the classification benefit of epoch-wise averaging remains uncertain, its use may still offer advantages over using a single epoch by reducing the influence of outliers or transient dynamics, potentially allowing to better capture stable diagnostic information, without an observed loss in performance. Studying the effect of varying the supporting epoch number further would be beneficial.

Second, we show that model choice is a determinant of classification accuracy. The best-performing classifiers (SVM-RBF, Multi-Layer Perceptron, k-Nearest Neighbours) assign a consistent label to 68.6% of the individuals, suggesting that the features likely contain diagnostic information allowing for multiple modelling approaches to succeed at classification. Notably, these are all models capable of learning nonlinear patterns, suggesting the relationships between networks features and diagnosis is likely complex. However, models vary in their ability to exploit diagnostic information; SVM-RBF demonstrates the strongest robustness to training set variations (65.7% receive a consistent classification across ten split iterations), with a comparable result for k-Nearest Neighbours (62.7%) Other models appear more sensitive to changes in the training data (Multi-Layer Perceptron consistency: 40.2%). Overall, although multiple nonlinear models provide a good chance at class separability, performance varies substantially, with some models representing the feature space more effectively than others.

Finally, feature selection and dimensionality reduction do not provide a significant advantage; the raw feature space was already appropriately low-dimensional (six predictors) and well-supported by the sample size.

Previous analyses of this dataset indicated that single-channel features were only informative when captured in proximity to epileptiform abnormalities.[12] Here, this subgroup was classified as accurately as those with a completely normal EEG examination and those with EEG captured in proximity to nonspecific abnormalities (65.8%, 66.3% and 71.5% respectively). This suggests network measures are robust to state-related variability, likely arising either from an intrinsic stability or from the attenuation of state-dependent information through epoch-wise averaging.

Results for network measures extracted from classical frequency bands show that, with all six features considered, low alpha achieves the highest balanced accuracies (51.7-65.8%), with delta and theta performing almost as well (balanced accuracies 48.5-60.2 across the two bands). Performance for alpha and beta is lower (40.5-52.8%). This suggests that network measures derived from lower frequency bands, and particularly low alpha (6-9 Hz), are more informative for this classification problem than those derived from higher frequency bands.

Our measures are selected based on evidence from the epilepsy literature and encode information on the strength of network coupling, the efficiency of information transfer, its local, hub, and hierarchical organisation, and its propensity for synchronisation. These might capture neurophysiologically meaningful cortical dynamics relevant to epilepsy, including pathological synchronisation, altered communication dynamics potentially facilitating synchronous firing, and a disproportionate influence of certain regions affecting network activity. Accordingly, these measures’ sensitivity is higher for our epilepsy sample, i.e., people with epilepsy are correctly classified more often than people with FDS (e.g., 78.4% and 53.1% correct classification respectively in our best configuration).

While these features show promise for supporting diagnostic decision-making, they may not be considered markers for a positive diagnosis of FDS. Achieving this would be crucial to allow improved mechanistic understanding and reduced stigma surrounding FDS. Whilst evidence to date remains limited regarding whether interictal EEG markers could support a positive diagnosis of FDS,[11,12,41–45] this continues to be an ongoing priority warranting further investigation. So far, the few studies with classification-based analyses avoiding data leakage suggest that single-channel measures might have limited potential.[12,41] Other connectivity-based or end-to-end approaches should be explored. Investments in design, sampling strategies and methodological quality will be essential for future advances.[11]

### Limitations, applicability, and future directions

As discussed in Faiman *et al.*[12], limitations include possible misdiagnosis for a minority of people without video-EEG confirmation, exclusion of people with unconfirmed diagnosis, and retrospective sampling. Applicability is limited to people without major neurological and psychiatric comorbidities. We do not exclude that network measures beyond those studied here might also prove informative.

This work involves further analyses of existing data, increasing the risk of Type I error. Testing multiple model configurations increases the likelihood that some will perform well by chance; however, the best-performing models are consistent across configurations and strongly surpass both permuation-based performance and models using confounders alone. As each outer fold may use different hyperparameter combinations, reported metrics reflect average model potential rather than performance of a single trained model.

As this study represents model development, findings require replication in independent samples using the most promising configurations, and validation across external centres to ensure generalisability. This study assessed whether quantitative interictal EEG analysis can provide supporting information for diagnostic decision-making; however, relevant demographic and clinical information could be integrated for further model gains. Ultimately, the clinical utility of any EEG-derived diagnostic models should be assessed in the context of existing clinical practice; real-world evaluations would help determine feasibility, with randomised clinical trials needed to establish whether improvements in diagnostic time and accuracy can be obtained and whether these translate to improved clinician confidence and patient outcomes.

## CONCLUSION

This diagnostic accuracy study assesses whether network measures derived from visually normal resting-state EEG can discriminate between a diagnosis of non-lesional epilepsy and one of functional/dissociative seizures (FDS). The balanced accuracy of multiple configurations tested achieve levels significantly higher than chance (best performing: 65.8%), establishing evidence for the clinical validity of these markers and highlighting their potential to support diagnostic probability estimation in the clinic. We offer recommendations on modelling strategies likely to be more effective for this diagnostic problem.

## DATA AVAILABILITY

Anonymised data are available upon request for collaborative purposes. The code written for this study is publicly available at [link to repository will be included once the manuscript is accepted].

## AUTHOR CONTRIBUTIONS

Conceptualisation: P.K., I.F., W.W., A.H.Y., P.S. Methodology: P.K., I.F., R.S., W.W., P.S. Investigation: I.F., N.C. Resources: J.S.W., F.B., P.S., A.H.Y. Data curation: P.K., I.F., N.C. Software and formal analyses (EEG): P.K., I.F. Software and formal analyses (statistical): P.K. Writing - original draft: P.K., I.F. Visualization: P.K. Writing - review and editing: P.K., W.W., R.S., J.S.W., F.B., N.C., A.H.Y., I.F., P.S. Supervision: W.W., R.S., J.S.W., F.B., A.H.Y, I.F., P.S. Funding acquisition: P.S., A.H.Y., W.W.

## ACKNOWLEDGEMENTS

We thank the personnel of King’s College Hospital Department of Neurophysiology for their work on EEG data acquisition and administrative support.

The computations described in this paper were performed using the University of Birmingham’s BlueBEAR HPC service, which provides a High Performance Computing Service to the University’s research community. See http://www.birmingham.ac.uk/bear for more details.

## FUNDING

I.F., P.S. and A.H.Y. acknowledge the support of the Bergqvist Charitable Trust through the Psychiatry Research Trust. P.K. acknowledges the support of the EPSRC (grant EP/V520275/1).

J.S.W receives support from the MRC Centre for Neurodevelopmental Disorders (MRC Grant Number MR/W006251/1)

The views expressed are those of the author(s) and not necessarily those of the funders, including NIHR or the Department of Health and Social Care.

## COMPETING INTERESTS

P.K., I.F., P.S., R.S., A.H.Y., N.C., F.B.: These authors report no competing interests.

W.W is co-founder, director and share-holder of Neuronostics.

J.S.W has received research support from Angelini Pharma and Jazz Pharmaceuticals, is the co-author of a patent regarding epileptic seizure prediction and a co-founder of NeuralPulse.

## SUPPLEMENTARY MATERIAL

See Supplementary Material file

## Notes

### Author Declarations

London Queen Square Research Ethics Committee of the NHS Health Research Authority gave ethical approval for this work.

### Summary of Updates

Improvements to statistical methodology (feature selection methods, permutation-based testing). Some passages from methods moved to supplemental file.

