## Supplementary Materials for "Resting-state EEG network markers differentiate people with epilepsy and functional seizures"

### SUPPLEMENTARY MATERIAL

**Section 4.1. Results for alternative model configurations: different feature selection approaches, dimensionality reduction methods and minimum number of EEG epochs** **18**

**Section 4.2. Results from the full feature set (six features, without feature selection)** .. **24**

#### Section 1. Additional details on sampling and participants

**Participant identification and matching procedure:**

In-length participant identification and matching information, including a patient flowchart, is provided in the Supplementary Material of Faiman et al., 2023, as cited in the main manuscript. In summary, eligible patients were identified through consecutive retrospective screening of specialist clinic lists and review of clinical notes relevant to their first presentation with suspected seizure disorder against inclusion/exclusion criteria. Initially, 161 eligible patients were identified. 106 people with epilepsy or FDS naturally matched by age at EEG (5-year range) and gender, resulting in 53 matched pairs. The remaining unmatched patients (18 females with FDS and 6 males with epilepsy), underwent targeted identification of matched counterparts using the Cogstack clinical information retrieval platform at King's College Hospital. Following matching, exclusion of people with an uncertain diagnosis (n=31), and exclusion of patients without retrievable EEG data (n=8), 146 patients with medication-free EEG recordings and a confirmed diagnosis were included (73 epilepsy, 73 FDS). By the time of data analysis, two additional patients who were initially classified as uncertain diagnosis received an established epilepsy diagnosis and were included in the analyses without a matched FDS counterpart.

**Details on inclusion and exclusion criteria:**

Exclusion criteria were acute symptomatic seizures, abnormal CT/MRI, eventual diagnosis of concurrent epilepsy and FDS, relevant history of other neurological, neurodevelopmental, or severe mental health disorders.

With regards to relevant history of other neurological disorders, this refers to disorders such as brain injury, ischaemic events, brain tumours, encephalitis, idiopathic intracranial hypertension, neural malformations, moderate to severe small vessel disease, cerebral hypoxia, neurodegenerative disorders, cerebral palsy, severe sleep disorders, brain surgery. We also excluded people with concurrent atrial fibrillation.

With regards to neurodevelopmental disorders, we excluded people with concurrent moderate to severe autism spectrum disorder, ADHD, or learning disabilities.

With regards to severe mental health disorders, we excluded people with active and severe mental health disorder such as major depressive disorder, bipolar disorder, active suicidal ideation, psychosis, schizophrenia, obsessive-compulsive disorder, addiction, multiple personality disorders, PTSD.

**Examples of concurrent disorders that did not lead to participant exclusion** are migraines, headaches, mild to moderate anxiety or depression, borderline personality disorder, mental health illness considered treated or resolved (e.g., past PTSD, depression, or self-harm), mild autism spectrum disorder, mild ADHD, dyslexia.

#### Section 2. Detailed methods

##### 2.1 Obtaining Networks

For computing functional networks, the 21 EEG channels corresponding to electrodes to be represented by nodes in the network are bandpass filtered to 6 – 9 Hz (low alpha range)[1–6] using a 4th-order Butterworth filter. Although spanning mainly the classical theta band, the 6 – 9 Hz range is referred to as low alpha[7] to maintain consistency with previous studies.[1–6] A Hilbert transform is applied. We then remove the first and last second from each 22-second epoch, to mitigate edge artifacts from the filtering and Hilbert transform. From each resulting 20-second filtered epoch, we calculate, pairwise between electrodes, the phase-locking factor (PLF)[8]:

$$PLF_{ij} = \left| \frac{1}{N} \sum_{n=1}^{N} e^{i\Delta \quad ij(t_n)} \right|, \qquad (1)$$

and lag[9]:

$$\tau_{ij} = arg\left( \sum_{n=1}^{N} e^{i\Delta \quad ij(t_n)} \right), \qquad (2)$$

where Δϕij(t) is the phase difference between signals i and j at time t. The PLF quantifies the consistency of phase difference between two signals over time; the lag reflects the direction and magnitude of that difference.

To establish a null distribution for PLF values, we generate 99 surrogate epochs, preserving power spectrum and autocorrelations but eliminating pairwise correlations, using a maximum of 100 iterations of the complex iterative amplitude-adjusted Fourier transform (iAAFT) algorithm.[10]

For each surrogate epoch, we calculate pairwise PLF values. To construct a network from the epoch PLF values, edges are prohibited if the PLF between the signals at the respective electrodes does not exceed that of 95% of the surrogate PLFs for the same pair. This retains only statistically significant connections. For each electrode pair (i, j) the respective candidate edge is considered to be directed from i to j, where the lag $\tau_{ij} > 0$.

From the remaining candidate edges i → j, we prohibit those for which a path exists of two or three edges from i to j, where all edges on this path have a PLF greater than that between i and j themselves, as this correlation can be better explained by the longer, indirect paths.[3] The remaining edges form the functional network, and are ascribed weights equal to

$$w_{ij} = \frac{PLF_{ij}}{\frac{1}{n}\sum_{i,j} A_{ij} PLF_{ij}}, \qquad (3)$$

where A is the unweighted adjacency matrix such that $A_{ij} = 1$ if there exist an edge i → j, zero otherwise. The division by $\frac{1}{n}\sum_{i,j} A_{ij}\mathrm{PLF}_{ij}$ performs a normalisation which sets the mean node strength equal to 1, allowing comparability of network structures across epochs and participants, disambiguated from a notion of global coupling strength.

Analyses for this project are conducted in Python 3.11.3 using the MNE,[11] numpy,[33] scipy,[13] numba,[14] NetworkX,[15], brain connectivity toolbox[16,17], trophic analysis toolbox[18], pandas[19,20], pycircstat[21], scikit-learn[22], imbalanced-learn[23], xgboost[24] and statsmodels[25] packages, alongside custom code. Supplementary Fig. S1 was generated using the seaborn package[26].

#### 2.2 Connectivity-based Measures

We calculate six functional connectivity-based measures covering a range of network properties selected based on previous evidence from the epilepsy literature.[1–3,6,27–34]

Whilst one measure, mean functional connectivity, is derived from the raw PLF values irrespective of the network structure, for the remaining measures we use the weighted network after edge rejection. We call the network G, for graph. Since each individual has several epochs of EEG, we test classifiers using the epoch-wise mean of each measure for each person.

##### 2.2.1 Mean Clustering Coefficient

Clustering is a topological network feature describing the tendency of connected nodes to have other connected nodes in common. This provides a measure of how locally interconnected neighbouring nodes are and is summarised by the mean local clustering coefficient, defined as

$$C(G) = \frac{1}{N}\sum_{i\in V}\frac{[(M+M^T)^3]_{ii}}{2\left[\left(d_i^{in}+d_i^{out}\right)\left(d_i^{in}+d_i^{out}-1\right)-2d_i^{\leftrightarrow}\right]}, \qquad (4)$$

where the (*i,j*)-th entry of the matrix *M* is the cube root of the associated edge-weight for a weighted network, and $d_i$ , $d_i^{out} \wedge d_i^{\leftrightarrow}$ are the number of in-edges, out-edges and bidirectional edges connected to node *i* respectively [35].

##### 2.2.2 Mean Functional Connectivity

To summarise the global strength of coupling in the network, we calculate mean functional connectivity, the average value of the PLF between all pairs of nodes, including those rejected as edges in the network

$$\mathrm{MFC} = \frac{1}{n(n-1)}\sum_{i\neq j}\mathrm{PLF}_{ij}. \qquad (5)$$

Note that the choice of normalisation factor in eq. 3 is introduced in order to disambiguate global coupling strength from the edge weights, significantly reducing correlations between MFC and other network measures.

##### 2.2.3 Network Efficiency

The shortest path length between two nodes represents how efficiently information can spread from one to the other. We represent this property globally with the network efficiency,[36]

$$E(G) = \frac{1}{n(n-1)} \sum_{i \neq j} \frac{1}{d_{ij}}, \qquad (6)$$

where dij is the shortest path-length from i to j, with the distance attributed to an edge counted as the inverse of the corresponding edge weight.

##### 2.2.4 Trophic Incoherence

We consider the notion of directionality as a property of the whole network, i.e., the extent to which a hierarchy exists in a directed network through which the edges have a predominant direction. This is explored through the ecological notion of trophic levels: the hierarchical positioning of organisms in a food chain or web[37], and quantified by the trophic incoherence, described by MacKay *et al*[18]. A low trophic incoherence value indicates a more "directed" or "hierarchical" network.

##### 2.2.5 Mean Betweenness Centrality

We also include a notion of node centrality, summarising the importance of individual nodes in the network structure. Betweenness centrality describes the frequency with which a node is visited on the shortest paths connecting pairs of other nodes, as follows[38]:

$$g(v) = \sum_{i \neq v \neq j} \frac{\sigma_{ij}(v)}{\sigma_{ij}}, \qquad (7)$$

where σij is the total number of shortest paths between nodes i and j, and σij (v) is the number of shortest paths between nodes i and j which pass through node v. To summarise this property globally, we take the mean of g over all nodes.

##### 2.2.6 Critical Coupling

We choose a notion of criticality based on a Kuramoto oscillator model of the brain, which describes a system of coupled populations of oscillators wherein network structure plays a crucial role in the dynamic behaviour of seizure propagation.[3,39] The critical coupling of this system is defined as the minimal value of the global coupling strength parameter for which non-trivial synchronous behaviour emerges in one or more populations of oscillators. This model-based measure represents the theoretical ability of seizures to emerge in a simulated network. Its calculation from data-derived networks is described in Schmidt *et al.*,[3] with the local intra-population coupling strength estimated by the standard deviation of the EEG.[6]

#### 2.3 Nested Cross-Validation

Cross validation ensures that the entire dataset is utilised in evaluating model performance. Many machine learning models have multiple configurations – determined by internal *hyperparameters* – which can be chosen for training. Selecting optimal hyperparameter values is nontrivial. Model training should not be influenced by the data used to test it; to avoid such contamination, any data-driven choices affecting an instance of the model should be made using training data only. To this end, we use a nested cross-validation with a second tier of cross-validation within each fold's training stage, in which each hyperparameter set is cross-validated, and the best-performing is selected for testing on the corresponding outer-tier testing set. This ensures model evaluation is based on sensible hyperparameter choices for each fold, but keeping model training decisions blinded to the testing data.

The nested cross-validation framework is described in Fig. 1. We distinguish between *model selection* – what features are considered and which type of model is suitable – and *hyperparameters,* parameter choices that characterise how the model learns from the data. Model selection includes model type, which dimensionality reduction method is used, and minimum number of EEG epochs required for inclusion in average feature estimation (Table 1). Hyperparameters include both pre-model operations (data transformation calculation and varying number of components/dimensions for dimensionality reduction), and model-specific hyperparameters considered during training (Table 1).

##### 2.3.1 Model Selection

**Minimum number of EEG epochs for average feature estimation.** To identify robust strategies for translational application, we test analytical recommendations to improve feature stability.[41] Since each individual has a varying number of EEG epochs from which mean network measures are derived, we assess whether averaging from a greater number of epochs provides more stable estimates, translating in improved classifier performance. We therefore run our analyses on both the full dataset, and the subset of individuals with at least four EEG epochs available. This cut-off is chosen to capture a nontrivial effect of multiple epochs whilst retaining a sufficiently large and balanced data subset.

**Feature selection.** As classifiers may be sensitive to the inclusion of redundant or correlated predictors, eliminating analytically non-contributory features is best practice.[40] In addition to reporting results from the whole feature set (six features), we test a feature selection methodology performing nested cross-validated backward feature elimination as follows. For each of the 100 outer-tier folds (10-fold split x 10 repeats of fold selection), we obtain the inner-tier cross-validated balanced accuracy of a naïve Bayes classifier model. Using a backwards-selection framework, we repeat the inner tier of cross-validation with each feature in turn eliminated. Taking the model with the highest balanced accuracy between the original model and models with a single feature removed, we iteratively continue feature elimination until balanced accuracy no longer increases. The internal cross-validated hyperparameter grid search is then implemented on the same development set with the selected reduced feature set. Testing is then performed on the hold-out set.

**Dimensionality reduction approach.** We test configurations with no dimensionality reduction, alongside two dimensionality reduction methods: principal component analysis (PCA),[42] a common linear technique, and spectral embedding,[43] a nonlinear technique to transform the data using mathematical properties of a matrix representation of the proximity of data points. The use of dimensionality reduction is justified by the possibility that this may uncover underlying patterns which are latent in the raw features.

**Model type.** We implement the cross-validation framework on 14 classification model types (Table 1). Support vector machine (SVM) classifiers with different kernel functions are treated as separate types as these models classify data by sufficiently distinct mechanisms.

##### 2.3.2 Hyperparameters

**Pre-model operations: data transformation approach.** As some machine learning models are assumed to perform better on normally-distributed data, the use of a Box-Cox transform[44] to approximate the training data to a normal distribution is implemented as a hyperparameter, along with the precision of the maximum likelihood estimation for the Box-Cox parameter, if applied. Because a full grid-search over a large number of Box-Cox parameters significantly increases computational expense, we limit these to three cases: no Box-Cox transform, and optimised Box-Cox parameter to zero or one decimal place(s).

**Pre-model operations: number of dimensions.** If the model selection tested includes dimensionality reduction, the number of components/dimensions (from one to four) is treated as a hyperparameter.

**Model-specific hyperparameters**. These are listed in Table 1.

##### 2.3.3 Nested cross-validation process

A model configuration consisting of a model type, dimensionality reduction approach and dataset (based on the inclusion of individuals with a minimum of one or four EEG epochs) is selected. Individuals are partitioned into ten independent folds stratified by diagnosis. Each fold is treated as a hold-out testing set with the complementary subset taken as development set. Within each development set, a further stratified ten-fold split is implemented, partitioning the data into ten non-overlapping testing sets with complementary training sets for hyperparameter tuning and model fitting.

Each training set is evaluated against all hyperparameter combinations, implementing a grid-search to optimise hyperparameters; the data are transformed using a Box-Cox transform (if applied), then whitened and, if relevant to the model configuration, transformed for dimensionality reduction with number of components from one to four. The model is then trained. The corresponding testing set is transformed using the optimal box-cox parameter, mean, standard deviation and, if applied, dimensionality reduction transform obtained from the training set, and the model is assessed on the transformed testing data. A weighted average of the balanced accuracy across all ten inner folds is calculated for each hyperparameter combination set; hyperparameters with the highest balanced accuracy are selected for evaluation on the hold-out testing set. Balanced accuracy ($\frac{1}{2}\left(\frac{TruePositives(epilepsy)}{Trueepilepsycases}+\frac{TruePositives(FDS)}{TrueFDScases}\right)$) is chosen as the outcome metric to equally represent the effect of correctly or incorrectly classifying samples in the epilepsy and FDS classes. The transformation and training process with the best-performing hyperparameters is then applied to the whole development set; the corresponding hold-out set is transformed accordingly and used to evaluate model performance.

The whole inner fold procedure is repeated for each of the ten development sets. We then obtain a weighted mean of balanced accuracy, as well as the weighted averages of sensitivity to the epilepsy class and sensitivity to the FDS class over the outer folds. To further improve generalisability, we repeat the process over ten iterations of random outer-fold selection and report means and standard deviations from these ten repeats in results tables.

The entire nested cross-validation framework is implemented for each model configuration tested, resulting in an overall balanced accuracy score which summarises the ability of a model configuration to classify our data, not associated to any specific hyperparameters. Random seeds are used for replicability of both inner- and outer-tier splits; however, note that since for each outer-fold we may train the model on a different set of optimal hyperparameters, we cannot claim that this score represents the classification accuracy of a specific model instance.

Within our differential diagnosis framework, we replace terminology of “sensitivity” and “specificity” with “sensitivity to the epilepsy class” and “sensitivity to the FDS class”, respectively. Conventionally, however, classification models require classes to be identified as positive and negative, respectively; we therefore code epilepsy as positive and FDS as negative.

##### 2.4 References for additional analyses section

Mann-Whitney U-test[45] is implemented to compare median average network measures between groups. A binomial generalised linear model (GLM) with a logit link function is fitted using Python statsmodels package[46] to ensure potential confounders (age, sex, presence of comorbidities) are not predictive of diagnosis in the subset with four or more epochs. Chi-squared test[47] is used to assess group differences in categorical variables (or Fisher's exact test[48] for categories containing fewer than five individuals); t-tests[49] for continuous normally-distributed variables, and Mann-Whitney U-tests[45] for continuous variables with other distributions. Watson-Williams test[50] is implemented to compare means of circular data (time of day of EEG recording, in radians).

#### Section 3. Feature selection results

In the full sample, backward feature elimination identifies a three-feature subset – network efficiency, trophic incoherence, and mean functional connectivity – as the most commonly-selected combination (14 of 100 outer iterations; Supplementary Table S1).

In the subset with at least four epochs, mean functional connectivity, network efficiency, critical coupling and betweenness centrality are the most commonly-selected combination (24 out of 100 iterations; Supplementary Table S2).

**Table S1.** Results of feature elimination on the 100 outer folds for the full sample. This table shows all feature sets which were selected by at least one fold, and the number of folds for which each was selected. Across all subsets tested during feature selection, network efficiency is selected most consistently (92 iterations), followed by trophic incoherence (81), mean functional connectivity (68), betweenness centrality (49), mean clustering coefficient (34), and critical coupling (34).

| Feature Set | Count |
|---|---|
| Network Efficiency, Trophic Incoherence, Mean Functional Connectivity | 14 |
| Network Efficiency, Trophic Incoherence, Betweenness Centrality | 10 |
| Network Efficiency, Trophic Incoherence | 10 |
| Mean Functional Connectivity, Network Efficiency, Trophic Incoherence, Critical Coupling | 8 |
| Mean Clustering Coefficient, Mean Functional Connectivity, Network Efficiency, Trophic Incoherence | 6 |
| Mean Clustering Coefficient, Mean Functional Connectivity, Network Efficiency, Trophic Incoherence, Critical Coupling | 6 |
| Mean Clustering Coefficient, Mean Functional Connectivity, Network Efficiency, Trophic Incoherence, Betweenness Centrality | 6 |
| Mean Clustering Coefficient, Mean Functional Connectivity, Network Efficiency, Critical Coupling, Betweenness Centrality | 6 |
| Mean Functional Connectivity, Network Efficiency, Trophic Incoherence, Betweenness Centrality | 5 |
| Mean Functional Connectivity, Network Efficiency, Trophic Incoherence, Critical Coupling, Betweenness Centrality | 3 |
| Network Efficiency, Betweenness Centrality | 3 |
| Network Efficiency, Trophic Incoherence, Critical Coupling, Betweenness Centrality | 3 |
| Mean Functional Connectivity, Trophic Incoherence, Betweenness Centrality | 3 |
| Mean Clustering Coefficient, Mean Functional Connectivity, Trophic Incoherence, Betweenness Centrality | 2 |
| Mean Clustering Coefficient, Mean Functional Connectivity, Network Efficiency, Betweenness Centrality | 2 |
| Mean Functional Connectivity, Network Efficiency, Critical Coupling, Betweenness Centrality | 2 |
| Mean Clustering Coefficient, Mean Functional Connectivity, Network Efficiency, Trophic Incoherence, Critical Coupling, Betweenness Centrality | 1 |
| Mean Functional Connectivity, Trophic Incoherence | 1 |
| Mean Clustering Coefficient, Network Efficiency, Trophic Incoherence, Betweenness Centrality | 1 |
| Mean Clustering Coefficient, Mean Functional Connectivity | 1 |
| Network Efficiency, Critical Coupling, Betweenness Centrality | 1 |
| Mean Functional Connectivity, Network Efficiency | 1 |
| Mean Clustering Coefficient, Network Efficiency, Critical Coupling, Betweenness Centrality | 1 |
| Mean Clustering Coefficient, Network Efficiency, Trophic Incoherence | 1 |
| Network Efficiency, Critical Coupling | 1 |
| Mean Clustering Coefficient, Mean Functional Connectivity, Critical Coupling | 1 |
| Network Efficiency, Trophic Incoherence, Critical Coupling | 1 |

**Table S2.** Results of feature elimination on the 100 outer folds for the subset with at least four epochs. This table shows all feature sets which were selected by at least one fold, and the number of folds for which each was selected. Across all subsets tested during feature selection, network efficiency is selected most consistently (96 iterations), followed by betweenness centrality (92), mean functional connectivity (70), critical coupling (57), mean clustering coefficient (28), and trophic incoherence (18).

| **Feature Set** | **Count** |
|---|---|
| Mean Functional Connectivity, Network Efficiency, Critical Coupling, Betweenness Centrality | 24 |
| Network Efficiency, Betweenness Centrality | 14 |
| Mean Functional Connectivity, Network Efficiency, Betweenness Centrality | 12 |
| Network Efficiency, Critical Coupling, Betweenness Centrality | 9 |
| Mean Clustering Coefficient, Mean Functional Connectivity, Network Efficiency, Betweenness Centrality | 8 |
| Mean Clustering Coefficient, Mean Functional Connectivity, Network Efficiency, Critical Coupling, Betweenness Centrality | 7 |
| Mean Clustering Coefficient, Mean Functional Connectivity, Network Efficiency, Trophic Incoherence, Critical Coupling, Betweenness Centrality | 4 |
| Mean Functional Connectivity, Network Efficiency, Trophic Incoherence, Critical Coupling, Betweenness Centrality | 3 |
| Mean Clustering Coefficient, Mean Functional Connectivity, Network Efficiency, Trophic Incoherence, Betweenness Centrality | 2 |
| Mean Clustering Coefficient, Mean Functional Connectivity, Network Efficiency, Trophic Incoherence, Critical Coupling | 2 |
| Mean Functional Connectivity, Network Efficiency | 2 |
| Mean Clustering Coefficient, Mean Functional Connectivity, Critical Coupling, Betweenness Centrality | 1 |
| Critical Coupling, Betweenness Centrality | 1 |
| Mean Clustering Coefficient, Network Efficiency, Trophic Incoherence, Critical Coupling | 1 |
| Mean Functional Connectivity | 1 |
| Network Efficiency, Trophic Incoherence, Critical Coupling, Betweenness Centrality | 1 |
| Mean Clustering Coefficient, Network Efficiency, Critical Coupling, Betweenness Centrality | 1 |
| Mean Functional Connectivity, Trophic Incoherence, Critical Coupling, Betweenness Centrality | 1 |
| Mean Functional Connectivity, Network Efficiency, Critical Coupling | 1 |
| Network Efficiency, Trophic Incoherence, Betweenness Centrality | 1 |
| Mean Functional Connectivity, Network Efficiency, Trophic Incoherence | 1 |
| Mean Functional Connectivity, Network Efficiency, Trophic Incoherence, Betweenness Centrality | 1 |
| Mean Clustering Coefficient, Network Efficiency, Trophic Incoherence, Critical Coupling, Betweenness Centrality | 1 |
| Mean Clustering Coefficient, Network Efficiency, Betweenness Centrality | 1 |

### Section 4. Additional cross-validation results

#### Section 4.1. Results for alternative model configurations: different feature selection approaches, dimensionality reduction methods and minimum number of EEG epochs

**Table S3**. Results of nested cross-validation for the full sample with feature selection and no dimensionality reduction. For each of the ten repeats of fold selection, each individual is classified exactly once as part of one of the ten non-overlapping testing folds. Means and standard deviations are calculated for outcome measures across these ten repeats. In addition, the 95$^{th}$ percentile mean balanced accuracy obtained from repeating the same nested cross-validation process for each model type on 99 permutations of the true classification labels is reported in the final column.

| Model | Balanced Accuracy mean (SD) (%) | Sensitivity (Epilepsy) mean (SD) (%) | Sensitivity (FDS) mean (SD) (%) | 95% Permutation Balanced Accuracy Quantile (%) |
|---|---|---|---|---|
| k-Nearest Neighbours | 63.4 (4.0) | 70.4 (5.8) | 56.4 (4.9) | 56.3 |
| RUSBoost | 62.3 (3.6) | 65.3 (3.6) | 59.3 (4.3) | 55.6 |
| Random Forest | 60.2 (3.4) | 63.5 (6.5) | 56.8 (4.1) | 57.5 |
| Support Vector Machine (RBF kernel) | 59.8 (4.1) | 65.2 (3.9) | 54.4 (6.1) | 56.9 |
| AdaBoost | 58.4 (2.9) | 62.4 (3.0) | 54.4 (4.6) | 55.9 |
| Multi-layer Perceptron | 57.4 (3.9) | 57.3 (5.6) | 57.4 (4.2) | 54.7 |
| XGBoost | 57.0 (2.9) | 59.5 (3.7) | 54.5 (5.2) | 57.0 |
| Decision Tree | 55.0 (4.7) | 57.6 (6.2) | 52.5 (4.4) | 57.0 |
| Linear Discriminant Analysis | 54.9 (5.1) | 60.7 (6.8) | 49.2 (5.5) | 57.9 |
| Gaussian Naive Bayes | 54.9 (4.2) | 72.7 (7.0) | 37.1 (5.8) | 56.2 |
| Support Vector Machine (Linear kernel) | 53.9 (3.6) | 68.0 (4.8) | 39.9 (5.4) | 58.4 |
| Logistic Regression | 53.9 (4.4) | 60.1 (6.6) | 47.7 (5.6) | 58.6 |
| Support Vector Machine (Sigmoid kernel) | 53.0 (5.0) | 60.1 (6.4) | 45.9 (8.8) | 55.1 |
| Dummy Classifier | 51.7 (3.6) | 68.4 (5.2) | 34.9 (5.4) | 52.6 |

**Table S4.** Results of nested cross-validation for the full sample with feature selection and principal component analysis dimensionality reduction. For each of the ten repeats of fold selection, each individual is classified exactly once as part of one of the ten non-overlapping testing folds. Means and standard deviations are calculated for outcome measures across these ten repeats. In addition, the 95th percentile mean balanced accuracy obtained from repeating the same nested cross-validation process for each model type on 99 permutations of the true classification labels is reported in the final column.

| Model | Balanced Accuracy mean (SD) (%) | Sensitivity (Epilepsy) mean (SD) (%) | Sensitivity (FDS) mean (SD) (%) | 95% Permutation Balanced Accuracy Quantile (%) |
|---|---|---|---|---|
| k-Nearest Neighbours | 61.8 (3.7) | 70.0 (5.4) | 53.7 (5.3) | 56.0 |
| RUSBoost | 61.2 (4.5) | 63.9 (6.5) | 58.5 (5.9) | 54.4 |
| Random Forest | 60.2 (3.3) | 62.4 (5.1) | 57.9 (3.6) | 56.7 |
| Support Vector Machine (RBF kernel) | 58.7 (5.1) | 63.5 (6.7) | 54.0 (5.2) | 55.7 |
| Multi-layer Perceptron | 57.9 (5.5) | 61.1 (8.4) | 54.7 (5.5) | 54.2 |
| XGBoost | 55.5 (3.1) | 56.9 (3.6) | 54.1 (5.2) | 54.8 |
| Gaussian Naive Bayes | 55.0 (3.7) | 72.1 (5.7) | 37.9 (5.9) | 57.2 |
| AdaBoost | 55.0 (2.1) | 58.3 (5.0) | 51.6 (3.1) | 55.2 |
| Decision Tree | 54.8 (4.0) | 56.7 (5.1) | 53.0 (5.5) | 55.0 |
| Logistic Regression | 53.4 (5.3) | 60.8 (7.7) | 46.0 (7.4) | 57.9 |
| Linear Discriminant Analysis | 53.3 (5.1) | 61.1 (7.7) | 45.6 (7.1) | 58.2 |
| Support Vector Machine (Linear kernel) | 53.2 (4.8) | 70.1 (5.9) | 36.3 (7.4) | 56.7 |
| Support Vector Machine (Sigmoid kernel) | 51.8 (3.6) | 56.3 (6.3) | 47.3 (4.1) | 53.8 |
| Dummy Classifier | 51.7 (3.6) | 68.4 (5.2) | 34.9 (5.4) | 52.0 |

**Table S5.** Results of nested cross-validation for the full sample with feature selection and spectral embedding dimensionality reduction. For each of the ten repeats of fold selection, each individual is classified exactly once as part of one of the ten non-overlapping testing folds. Means and standard deviations are calculated for outcome measures across these ten repeats. In addition, the 95$^{th}$ percentile mean balanced accuracy obtained from repeating the same nested cross-validation process for each model type on 99 permutations of the true classification labels is reported in the final column.

| Model | Balanced Accuracy mean (SD) (%) | Sensitivity (Epilepsy) mean (SD) (%) | Sensitivity (FDS) mean (SD) (%) | 95% Permutation Balanced Accuracy Quantile (%) |
|---|---|---|---|---|
| Support Vector Machine (RBF kernel) | 60.0 (3.7) | 75.6 (3.7) | 44.4 (5.8) | 55.7 |
| Random Forest | 58.9 (3.6) | 63.9 (4.3) | 53.8 (3.5) | 55.1 |
| k-Nearest Neighbours | 58.4 (4.1) | 69.2 (4.5) | 47.5 (4.9) | 55.4 |
| AdaBoost | 57.4 (2.7) | 59.9 (4.5) | 54.9 (3.7) | 54.1 |
| XGBoost | 57.1 (3.4) | 58.7 (4.6) | 55.6 (5.0) | 54.3 |
| Decision Tree | 56.7 (2.9) | 65.3 (4.4) | 48.1 (3.4) | 54.8 |
| RUSBoost | 55.8 (4.7) | 57.6 (6.8) | 54.0 (4.8) | 54.1 |
| Gaussian Naive Bayes | 55.6 (3.0) | 86.8 (3.8) | 24.4 (4.5) | 54.5 |
| Multi-layer Perceptron | 54.5 (2.9) | 70.8 (5.5) | 38.2 (5.7) | 55.9 |
| Linear Discriminant Analysis | 52.1 (3.3) | 80.9 (5.3) | 23.3 (3.9) | 56.1 |
| Logistic Regression | 51.8 (4.3) | 80.7 (7.1) | 23.0 (4.5) | 56.1 |
| Dummy Classifier | 51.7 (3.6) | 68.4 (5.2) | 34.9 (5.4) | 52.2 |
| Support Vector Machine (Sigmoid kernel) | 51.5 (2.8) | 68.9 (4.9) | 34.0 (5.1) | 54.7 |
| Support Vector Machine (Linear kernel) | 50.0 (0.0) | 100.0 (0.0) | 0.0 (0.0) | 50.1 |

**Table S6**. Results of nested cross-validation for the subset with a minimum 4 epochs per person with feature selection and no dimensionality reduction. For each of the ten repeats of fold selection, each individual is classified exactly once as part of one of the ten non-overlapping testing folds. Means and standard deviations are calculated for outcome measures across these ten repeats. In addition, the 95th percentile mean balanced accuracy obtained from repeating the same nested cross-validation process for each model type on 99 permutations of the true classification labels is reported in the final column.

| Model | Balanced Accuracy mean (std) (%) | Sensitivity (Epilepsy) mean (std) (%) | Sensitivity (FDS) mean (std) (%) | 95% Permutation Balanced Accuracy Quantile (%) |
|---|---|---|---|---|
| Support Vector Machine (RBF kernel) | 64.1 (3.0) | 78.6 (4.8) | 49.6 (5.3) | 57.1 |
| k-Nearest Neighbours | 64.0 (2.5) | 75.6 (6.0) | 52.4 (3.5) | 57.3 |
| RUSBoost | 61.4 (4.1) | 67.2 (4.1) | 55.6 (7.0) | 57.5 |
| XGBoost | 61.0 (2.3) | 70.7 (3.8) | 51.3 (5.7) | 58.9 |
| Random Forest | 60.9 (3.1) | 74.6 (6.8) | 47.3 (6.8) | 57.3 |
| AdaBoost | 60.5 (3.2) | 69.1 (3.2) | 51.8 (6.0) | 57.2 |
| Multi-layer Perceptron | 60.0 (4.2) | 62.8 (4.9) | 57.1 (6.5) | 57.4 |
| Decision Tree | 58.3 (5.5) | 72.5 (5.7) | 44.2 (7.5) | 57.1 |
| Support Vector Machine (Sigmoid kernel) | 56.1 (4.3) | 69.8 (6.5) | 42.4 (6.3) | 56.0 |
| Gaussian Naive Bayes | 54.9 (5.1) | 71.9 (6.7) | 37.8 (6.0) | 58.7 |
| Linear Discriminant Analysis | 54.3 (3.6) | 71.2 (6.6) | 37.3 (5.3) | 58.8 |
| Logistic Regression | 53.6 (4.5) | 71.1 (7.2) | 36.2 (7.1) | 58.8 |
| Dummy Classifier | 52.1 (4.9) | 68.1 (4.7) | 36.2 (7.7) | 54.2 |
| Support Vector Machine (Linear kernel) | 51.0 (2.9) | 78.1 (6.2) | 24.0 (4.3) | 58.8 |

**Table S7.** Results of nested cross-validation for the subset with a minimum 4 epochs per person, with feature selection and PCA dimensionality reduction. For each of the ten repeats of fold selection, each individual is classified exactly once as part of one of the ten non-overlapping testing folds. Means and standard deviations are calculated for outcome measures across these ten repeats. In addition, the 95th percentile mean balanced accuracy obtained from repeating the same nested cross-validation process for each model type on 99 permutations of the true classification labels is reported in the final column.

| Model | Balanced Accuracy mean (SD) (%) | Sensitivity (Epilepsy) mean (SD) (%) | Sensitivity (FDS) mean (SD) (%) | 95% Permutation Balanced Accuracy Quantile (%) |
|---|---|---|---|---|
| Support Vector Machine (RBF kernel) | 62.9 (3.7) | 76.5 (5.0) | 49.3 (6.7) | 55.6 |
| k-Nearest Neighbours | 62.2 (3.1) | 73.3 (6.2) | 51.1 (3.6) | 57.1 |
| Multi-layer Perceptron | 59.3 (2.8) | 61.9 (5.1) | 56.7 (5.6) | 57.0 |
| RUSBoost | 58.5 (4.6) | 63.5 (5.9) | 53.6 (4.9) | 58.1 |
| Random Forest | 57.9 (3.0) | 70.2 (6.0) | 45.6 (5.0) | 57.7 |
| AdaBoost | 55.6 (3.6) | 63.3 (5.3) | 47.8 (6.3) | 55.8 |
| XGBoost | 55.1 (3.5) | 62.8 (4.7) | 47.3 (5.2) | 56.7 |
| Gaussian Naive Bayes | 53.0 (4.7) | 73.3 (4.9) | 32.7 (6.9) | 57.2 |
| Decision Tree | 52.8 (3.6) | 67.0 (4.6) | 38.7 (7.4) | 55.3 |
| Logistic Regression | 52.5 (5.0) | 79.3 (6.5) | 25.8 (6.2) | 56.8 |
| Linear Discriminant Analysis | 52.3 (5.4) | 79.3 (7.0) | 25.3 (7.2) | 57.4 |
| Dummy Classifier | 52.1 (4.9) | 68.1 (4.7) | 36.2 (7.7) | 53.7 |
| Support Vector Machine (Sigmoid kernel) | 51.9 (4.2) | 59.8 (7.0) | 44.0 (6.1) | 53.9 |
| Support Vector Machine (Linear kernel) | 50.1 (3.3) | 79.1 (6.5) | 21.1 (5.6) | 56.5 |

**Table S8.** Results of nested cross-validation for the subset with a minimum 4 epochs per person, with feature selection and spectral embedding dimensionality reduction. For each of the ten repeats of fold selection, each individual is classified exactly once as part of one of the ten non-overlapping testing folds. Means and standard deviations are calculated for outcome measures across these ten repeats. In addition, the 95$^{th}$ percentile mean balanced accuracy obtained from repeating the same nested cross-validation process for each model type on 99 permutations of the true classification labels is reported in the final column.

| **Model** | **Balanced Accuracy mean (SD) (%)** | **Sensitivity (Epilepsy) mean (SD) (%)** | **Sensitivity (FDS) mean (SD) (%)** | **95% Permutation Balanced Accuracy Quantile (%)** |
|---|---|---|---|---|
| k-Nearest Neighbours | 58.5 (4.5) | 72.1 (3.6) | 44.9 (7.9) | 54.4 |
| Support Vector Machine (RBF kernel) | 57.4 (3.7) | 83.9 (4.5) | 30.9 (4.6) | 55.1 |
| XGBoost | 56.8 (4.2) | 64.2 (4.9) | 49.3 (5.8) | 54.4 |
| RUSBoost | 56.2 (4.7) | 58.8 (5.7) | 53.6 (6.1) | 55.5 |
| AdaBoost | 56.0 (2.9) | 65.1 (4.3) | 46.9 (6.0) | 55.2 |
| Random Forest | 55.8 (2.4) | 71.6 (4.8) | 40.0 (4.6) | 55.8 |
| Multi-layer Perceptron | 55.7 (5.3) | 74.2 (5.6) | 37.1 (8.9) | 54.6 |
| Decision Tree | 54.3 (2.3) | 64.2 (4.9) | 44.4 (4.3) | 55.4 |
| Logistic Regression | 52.6 (4.1) | 81.9 (3.7) | 23.3 (6.4) | 56.3 |
| Gaussian Naive Bayes | 52.4 (3.4) | 86.5 (2.7) | 18.2 (5.0) | 54.2 |
| Dummy Classifier | 52.1 (4.9) | 68.1 (4.7) | 36.2 (7.7) | 53.6 |
| Linear Discriminant Analysis | 51.8 (5.5) | 81.6 (4.3) | 22.0 (8.5) | 56.9 |
| Support Vector Machine (Linear kernel) | 50.6 (1.3) | 99.5 (0.8) | 1.8 (2.6) | 50.2 |
| Support Vector Machine (Sigmoid kernel) | 48.7 (3.0) | 70.5 (5.9) | 26.9 (2.1) | 52.6 |

#### Section 4.2. Results from the full feature set (six features, without feature selection)

**Table S9.** Results of nested cross-validation for the full sample and no dimensionality reduction, with all six features. For each of the ten repeats of fold selection, each individual is classified exactly once as part of one of the ten non-overlapping testing folds. Means and standard deviations are calculated for outcome measures across these ten repeats. In addition, the 95$^{th}$ percentile mean balanced accuracy obtained from repeating the same nested cross-validation process for each model type on 99 permutations of the true classification labels is reported in the final column.

| Model | Balanced Accuracy mean (SD) (%) | Sensitivity (Epilepsy) mean (SD) (%) | Sensitivity (FDS) mean (SD) (%) | 95% Permutation Balanced Accuracy Quantile (%) |
|---|---|---|---|---|
| RUSBoost | 62.9 (4.0) | 66.4 (4.4) | 59.3 (5.3) | 56.6 |
| k-Nearest Neighbours | 62.0 (2.5) | 69.3 (4.5) | 54.7 (4.1) | 56.1 |
| Support Vector Machine (RBF kernel) | 60.7 (4.5) | 66.4 (3.9) | 55.1 (5.8) | 56.3 |
| Random Forest | 60.6 (3.2) | 64.4 (4.4) | 56.8 (3.6) | 57.5 |
| Multi-layer Perceptron | 60.6 (3.7) | 61.2 (4.3) | 60.0 (4.6) | 56.2 |
| AdaBoost | 59.1 (3.3) | 61.9 (4.8) | 56.3 (4.8) | 56.5 |
| XGBoost | 58.1 (3.3) | 60.8 (3.8) | 55.3 (5.6) | 56.5 |
| Decision Tree | 56.1 (4.1) | 61.6 (6.2) | 50.7 (3.8) | 58.1 |
| Linear Discriminant Analysis | 56.1 (3.7) | 60.9 (5.3) | 51.2 (4.1) | 55.9 |
| Logistic Regression | 54.7 (3.6) | 59.7 (4.6) | 49.6 (4.1) | 56.5 |
| Support Vector Machine (Sigmoid kernel) | 54.1 (5.1) | 58.5 (6.3) | 49.6 (5.8) | 56.1 |
| Support Vector Machine (Linear kernel) | 53.9 (4.4) | 65.2 (5.3) | 42.6 (5.8) | 57.4 |
| Gaussian Naive Bayes | 53.7 (4.2) | 69.2 (7.5) | 38.2 (6.2) | 57.5 |
| Dummy Classifier | 51.7 (3.6) | 68.4 (5.2) | 34.9 (5.4) | 51.9 |

**Table S10.** Results of nested cross-validation for the full sample and PCA dimensionality reduction, with all six features. For each of the ten repeats of fold selection, each individual is classified exactly once as part of one of the ten non-overlapping testing folds. Means and standard deviations are calculated for outcome measures across these ten repeats. In addition, the 95$^{th}$ percentile mean balanced accuracy obtained from repeating the same nested cross-validation process for each model type on 99 permutations of the true classification labels is reported in the final column.

| Model | Balanced Accuracy mean (SD) (%) | Sensitivity (Epilepsy) mean (SD) (%) | Sensitivity (FDS) mean (SD) (%) | 95% Permutation Balanced Accuracy Quantile (%) |
|---|---|---|---|---|
| k-Nearest Neighbours | 61.5 (2.9) | 68.1 (3.8) | 54.9 (4.6) | 56.6 |
| Random Forest | 60.8 (2.8) | 63.5 (3.5) | 58.1 (3.4) | 57.8 |
| RUSBoost | 58.9 (3.3) | 61.9 (4.1) | 56.0 (3.3) | 56.3 |
| Multi-layer Perceptron | 58.7 (3.8) | 59.6 (5.0) | 57.8 (4.5) | 56.1 |
| AdaBoost | 58.1 (2.8) | 60.5 (4.1) | 55.8 (4.0) | 57.3 |
| Decision Tree | 58.1 (4.6) | 62.0 (4.7) | 54.1 (6.7) | 54.6 |
| Support Vector Machine (RBF kernel) | 57.9 (3.5) | 63.1 (2.0) | 52.7 (6.9) | 57.8 |
| XGBoost | 57.9 (2.6) | 59.2 (4.9) | 56.6 (2.7) | 56.1 |
| Linear Discriminant Analysis | 53.5 (4.1) | 59.5 (6.7) | 47.5 (5.3) | 58.7 |
| Logistic Regression | 53.3 (4.5) | 60.5 (7.4) | 46.2 (4.8) | 58.3 |
| Support Vector Machine (Sigmoid kernel) | 53.3 (4.6) | 56.7 (5.4) | 49.9 (4.8) | 53.6 |
| Gaussian Naive Bayes | 52.3 (3.5) | 68.4 (6.3) | 36.2 (5.7) | 57.8 |
| Dummy Classifier | 51.7 (3.6) | 68.4 (5.2) | 34.9 (5.4) | 51.9 |
| Support Vector Machine (Linear kernel) | 51.4 (5.2) | 64.7 (6.9) | 38.2 (5.4) | 57.5 |

**Table S11.** Results of nested cross-validation for the full sampleand spectral embedding dimensionality reduction, with all six features. For each of the ten repeats of fold selection, each individual is classified exactly once as part of one of the ten non-overlapping testing folds. Means and standard deviations are calculated for outcome measures across these ten repeats. In addition, the 95th percentile mean balanced accuracy obtained from repeating the same nested cross-validation process for each model type on 99 permutations of the true classification labels is reported in the final column.

| Model | Balanced Accuracy mean (SD) (%) | Sensitivity (Epilepsy) mean (SD) (%) | Sensitivity (FDS) mean (SD) (%) | 95% Permutation Balanced Accuracy Quantile (%) |
|---|---|---|---|---|
| Support Vector Machine (RBF kernel) | 62.0 (3.2) | 71.9 (5.4) | 52.1 (5.5) | 55.5 |
| AdaBoost | 61.0 (3.6) | 64.3 (7.4) | 57.8 (2.9) | 54.6 |
| Decision Tree | 60.9 (4.7) | 67.3 (5.9) | 54.4 (5.3) | 54.2 |
| Random Forest | 60.8 (4.6) | 64.5 (6.6) | 57.0 (4.6) | 55.7 |
| k-Nearest Neighbours | 60.3 (2.4) | 68.4 (5.1) | 52.2 (4.9) | 56.0 |
| XGBoost | 60.2 (3.7) | 62.9 (5.0) | 57.4 (4.6) | 54.7 |
| RUSBoost | 59.0 (5.1) | 58.9 (7.7) | 59.0 (4.1) | 54.9 |
| Multi-layer Perceptron | 56.0 (2.7) | 63.9 (6.7) | 48.2 (5.3) | 52.9 |
| Gaussian Naive Bayes | 53.8 (3.7) | 86.1 (2.8) | 21.5 (5.8) | 53.4 |
| Support Vector Machine (Sigmoid kernel) | 53.1 (2.8) | 66.9 (4.3) | 39.2 (5.4) | 52.6 |
| Linear Discriminant Analysis | 52.7 (4.0) | 80.3 (6.0) | 25.1 (4.2) | 54.1 |
| Dummy Classifier | 51.7 (3.6) | 68.4 (5.2) | 34.9 (5.4) | 52.0 |
| Logistic Regression | 50.9 (2.7) | 81.1 (4.2) | 20.7 (2.9) | 53.3 |
| Support Vector Machine (Linear kernel) | 50.3 (0.3) | 100.0 (0.0) | 0.5 (0.7) | 50.2 |

**Table S12.** Results of nested cross-validation for the subset with a minimum 4 epochs per person and PCA dimensionality reduction, with all six features. For each of the ten repeats of fold selection, each individual is classified exactly once as part of one of the ten non-overlapping testing folds. Means and standard deviations are calculated for outcome measures across these ten repeats. In addition, the 95th percentile mean balanced accuracy obtained from repeating the same nested cross-validation process for each model type on 99 permutations of the true classification labels is reported in the final column.

| Model | Balanced Accuracy mean (SD) (%) | Sensitivity (Epilepsy) mean (SD) (%) | Sensitivity (FDS) mean (SD) (%) | 95% Permutation Balanced Accuracy Quantile (%) |
|---|---|---|---|---|
| Multi-layer Perceptron | 65.0 (4.1) | 66.3 (3.7) | 63.8 (7.4) | 58.3 |
| Support Vector Machine (RBF kernel) | 62.6 (4.1) | 74.6 (6.3) | 50.7 (6.3) | 56.4 |
| k-Nearest Neighbours | 62.3 (4.4) | 74.9 (7.7) | 49.8 (5.2) | 59.2 |
| Random Forest | 58.8 (3.0) | 72.5 (5.6) | 45.1 (5.4) | 57.5 |
| RUSBoost | 58.4 (4.4) | 64.7 (5.5) | 52.0 (7.2) | 57.6 |
| AdaBoost | 57.2 (3.0) | 67.0 (6.5) | 47.3 (4.8) | 56.1 |
| XGBoost | 55.8 (4.3) | 66.8 (5.5) | 44.7 (5.3) | 57.2 |
| Support Vector Machine (Sigmoid kernel) | 54.8 (4.4) | 66.1 (5.7) | 43.6 (7.2) | 55.8 |
| Decision Tree | 54.1 (3.8) | 70.0 (5.6) | 38.2 (7.2) | 56.7 |
| Linear Discriminant Analysis | 52.5 (4.2) | 72.8 (7.7) | 32.2 (7.8) | 59.7 |
| Logistic Regression | 52.3 (3.7) | 72.6 (6.4) | 32.0 (9.0) | 61.0 |
| Dummy Classifier | 52.1 (4.9) | 68.1 (4.7) | 36.2 (7.7) | 53.2 |
| Gaussian Naive Bayes | 51.6 (3.8) | 71.4 (7.1) | 31.8 (5.7) | 59.2 |
| Support Vector Machine (Linear kernel) | 51.1 (4.1) | 74.7 (6.5) | 27.6 (5.8) | 60.0 |

**Table S13.** Results of nested cross-validation for the subset with a minimum 4 epochs per person and spectral embedding dimensionality reduction, with all six features. For each of the ten repeats of fold selection, each individual is classified exactly once as part of one of the ten non-overlapping testing folds. Means and standard deviations are calculated for outcome measures across these ten repeats. In addition, the 95$^{th}$ percentile mean balanced accuracy obtained from repeating the same nested cross-validation process for each model type on 99 permutations of the true classification labels is reported in the final column.

| Model | Balanced Accuracy mean (SD) (%) | Sensitivity (Epilepsy) mean (SD) (%) | Sensitivity (FDS) mean (SD) (%) | 95% Permutation Balanced Accuracy Quantile (%) |
|---|---|---|---|---|
| k-Nearest Neighbours | 60.9 (2.7) | 74.7 (5.2) | 47.1 (2.8) | 59.0 |
| Multi-layer Perceptron | 60.5 (2.5) | 72.8 (4.8) | 48.2 (6.9) | 57.4 |
| Support Vector Machine (RBF kernel) | 60.1 (2.2) | 78.6 (3.7) | 41.6 (3.6) | 58.8 |
| Random Forest | 59.3 (3.4) | 70.4 (5.8) | 48.2 (3.2) | 56.9 |
| AdaBoost | 58.1 (4.6) | 65.8 (6.4) | 50.4 (6.7) | 55.9 |
| XGBoost | 57.2 (5.9) | 63.9 (6.5) | 50.4 (9.2) | 56.2 |
| RUSBoost | 57.0 (5.2) | 61.8 (6.3) | 52.2 (6.3) | 57.8 |
| Decision Tree | 56.7 (4.5) | 63.7 (5.3) | 49.8 (7.2) | 55.2 |
| Logistic Regression | 54.7 (4.1) | 74.9 (4.7) | 34.4 (6.5) | 58.0 |
| Linear Discriminant Analysis | 53.8 (4.0) | 78.1 (4.2) | 29.6 (7.6) | 58.4 |
| Support Vector Machine (Linear kernel) | 53.4 (2.6) | 99.3 (0.9) | 7.6 (5.2) | 52.4 |
| Support Vector Machine (Sigmoid kernel) | 53.2 (4.8) | 70.2 (6.5) | 36.2 (5.8) | 55.0 |
| Dummy Classifier | 52.1 (4.9) | 68.1 (4.7) | 36.2 (7.7) | 53.9 |
| Gaussian Naive Bayes | 51.2 (3.2) | 81.1 (2.6) | 21.3 (5.9) | 57.7 |

**Table S14**. Subgroup analyses for the full sample and no dimensionality reduction, with all six features.

| Subgroup | Size of epilepsy subgroup | Size of FDS subgroup | Balanced Accuracy (%) | Sensitivity (Epilepsy) (%) | Sensitivity (FDS) (%) |
|---|---|---|---|---|---|
| **Sex, male** | 28 | 26 | 59.8 | 66.1 | 53.5 |
| **Sex, female** | 47 | 47 | 61.3 | 66.6 | 56.0 |
| **EEG normal** | 22 | 57 | 61.9 | 73.2 | 50.7 |
| **EEG abnormal, non-specific** | 25 | 14 | 66.4 | 59.2 | 73.6 |
| **EEG abnormal, epilepsy-specific** | 28 | 2 | 58.7 | 67.5 | 50.0 |
| **Comorbidity, absent** | 59 | 45 | 62.6 | 62.0 | 63.1 |
| **Comorbidity, present** | 16 | 28 | 62.3 | 82.5 | 42.1 |
| **Epilepsy type, focal** | 33 | 0 | n/a | 73.0 | n/a |
| **Epilepsy type, generalised** | 19 | 0 | n/a | 67.9 | n/a |
| **Epilepsy type, unknown** | 20 | 0 | n/a | 57.5 | n/a |
| **Functional seizure not captured during EEG appointment** | 0 | 43 | n/a | n/a | 57.0 |
| **Functional seizure captured during EEG appointment** | 0 | 30 | n/a | n/a | 52.3 |

#### Section 4.3. Results from classical frequency bands

The following are results for nested cross-validation on the six network metrics described in section 2.3, calculated using networks from EEG filtered to four classical frequency ranges: alpha (8-12.5 Hz), beta (13-30 Hz), delta (1-3.5 Hz) and theta (4-7.5 Hz)

**Table S15.** Results of nested cross-validation for the six features calculated on EEG filtered to delta band (1-3.5 Hz), with no dimensionality reduction. For each of the ten repeats of fold selection, each individual is classified exactly once as part of one of the ten non-overlapping testing folds. Means and standard deviations are calculated for outcome measures across these ten repeats.

| | Full Sample | | | | Subset (min. 4 epochs) | | |
|---|---|---|---|---|---|---|---|
| **Model** | **Balanced Accuracy mean (SD) (%)** | **Sensitivity (Epilepsy) mean (SD) (%)** | **Sensitivity (FDS) mean (SD) (%)** | | **Balanced Accuracy mean (SD) (%)** | **Sensitivity (Epilepsy) mean (SD) (%)** | **Sensitivity (FDS) mean (SD) (%)** |
| Decision Tree | 58.4 (3.5) | 70.1 (5.4) | 46.6 (4.0) | | 50.6 (4.5) | 50.0 (5.4) | 51.1 (8.8) |
| Random Forest | 56.5 (3.4) | 58.8 (4.7) | 54.1 (3.5) | | 55.4 (4.3) | 67.4 (3.4) | 43.3 (5.3) |
| Gaussian Naive Bayes | 56.1 (3.4) | 55.5 (5.5) | 56.7 (3.5) | | 58.2 (4.7) | 65.1 (7.3) | 51.3 (6.8) |
| AdaBoost | 55.1 (3.7) | 55.3 (6.1) | 54.9 (4.9) | | 50.1 (4.2) | 57.9 (7.2) | 42.2 (7.6) |
| RUSBoost | 53.0 (2.8) | 52.9 (5.1) | 53.0 (3.2) | | 50.7 (4.0) | 57.7 (4.4) | 43.8 (6.6) |
| k-Nearest Neighbours | 52.7 (4.3) | 44.4 (4.7) | 61.1 (5.2) | | 53.4 (4.0) | 57.0 (3.7) | 49.8 (6.2) |
| XGBoost | 51.9 (3.4) | 53.9 (6.9) | 50.0 (5.3) | | 51.0 (5.0) | 61.9 (3.9) | 40.0 (7.2) |
| Dummy Classifier | 51.7 (3.6) | 68.4 (5.2) | 34.9 (5.4) | | 52.1 (4.9) | 68.1 (4.7) | 36.2 (7.7) |
| Support Vector Machine (Linear kernel) | 51.0 (5.5) | 59.7 (7.4) | 42.3 (5.2) | | 50.4 (5.0) | 75.1 (5.9) | 25.8 (7.7) |
| Support Vector Machine (RBF kernel) | 50.6 (3.4) | 54.0 (5.1) | 47.3 (3.0) | | 53.1 (2.7) | 64.2 (3.5) | 42.0 (5.5) |
| Multi-layer Perceptron | 50.5 (3.0) | 47.1 (5.7) | 53.8 (2.5) | | 52.9 (5.7) | 50.5 (9.5) | 55.3 (6.6) |
| Logistic Regression | 49.9 (4.2) | 55.2 (5.0) | 44.7 (5.1) | | 50.6 (6.0) | 73.0 (7.2) | 28.2 (9.3) |
| Support Vector Machine (Sigmoid kernel) | 49.5 (3.3) | 52.5 (4.8) | 46.4 (6.9) | | 48.5 (5.9) | 59.8 (7.2) | 37.1 (8.2) |
| Linear Discriminant Analysis | 48.5 (4.6) | 54.1 (5.6) | 42.9 (6.2) | | 51.4 (6.4) | 71.8 (6.9) | 31.1 (10.0) |

**Table S16.** Results of nested cross-validation for the six features calculated on EEG filtered to theta band (4-7.5 Hz), with no dimensionality reduction. For each of the ten repeats of fold selection, each individual is classified exactly once as part of one of the ten non-overlapping testing folds. Means and standard deviations are calculated for outcome measures across these ten repeats.

| | Full Sample | | | | Subset (min. 4 epochs) | | |
|---|---|---|---|---|---|---|---|
| **Model** | **Balanced Accuracy mean (SD) (%)** | **Sensitivity (Epilepsy) mean (SD) (%)** | **Sensitivity (FDS) mean (SD) (%)** | | **Balanced Accuracy mean (SD) (%)** | **Sensitivity (Epilepsy) mean (SD) (%)** | **Sensitivity (FDS) mean (SD) (%)** |
| Linear Discriminant Analysis | 60.2 (2.5) | 64.8 (5.1) | 55.6 (5.2) | | 59.3 (5.2) | 76.0 (5.0) | 42.7 (7.6) |
| Support Vector Machine (RBF kernel) | 60.1 (3.8) | 68.4 (5.0) | 51.8 (5.5) | | 56.3 (6.5) | 72.5 (5.6) | 40.2 (8.8) |
| Support Vector Machine (Sigmoid kernel) | 59.9 (4.6) | 62.8 (5.7) | 57.0 (7.8) | | 56.7 (4.3) | 65.3 (7.5) | 48.2 (7.6) |
| Logistic Regression | 59.6 (3.8) | 64.1 (6.2) | 55.1 (5.2) | | 59.5 (6.0) | 78.9 (5.7) | 40.0 (9.4) |
| Gaussian Naive Bayes | 59.5 (3.8) | 69.9 (4.7) | 49.0 (6.4) | | 59.6 (7.9) | 71.4 (6.8) | 47.8 (10.8) |
| Support Vector Machine (Linear kernel) | 59.2 (2.9) | 66.8 (5.0) | 51.5 (5.9) | | 56.8 (6.6) | 75.1 (4.4) | 38.4 (10.2) |
| Multi-layer Perceptron | 58.0 (3.4) | 59.7 (6.2) | 56.3 (5.1) | | 56.8 (6.7) | 67.4 (5.0) | 46.2 (10.6) |
| XGBoost | 56.5 (4.7) | 54.1 (5.6) | 58.8 (7.2) | | 55.5 (4.1) | 60.2 (6.6) | 50.9 (5.4) |
| k-Nearest Neighbours | 56.3 (4.5) | 63.9 (4.4) | 48.8 (7.6) | | 54.9 (6.6) | 68.6 (6.4) | 41.1 (9.4) |
| Random Forest | 56.0 (3.3) | 61.5 (4.9) | 50.5 (4.9) | | 58.5 (6.3) | 68.1 (6.3) | 48.9 (7.7) |
| AdaBoost | 54.6 (4.3) | 56.3 (4.6) | 52.9 (6.3) | | 54.0 (3.6) | 60.2 (7.2) | 47.8 (7.4) |
| RUSBoost | 54.5 (3.4) | 54.5 (5.4) | 54.4 (5.3) | | 56.9 (5.2) | 54.4 (7.4) | 59.3 (5.7) |
| Dummy Classifier | 51.7 (3.6) | 68.4 (5.2) | 34.9 (5.4) | | 52.1 (4.9) | 68.1 (4.7) | 36.2 (7.7) |
| Decision Tree | 51.3 (3.7) | 52.1 (4.6) | 50.4 (5.3) | | 54.1 (4.8) | 59.8 (7.2) | 48.4 (9.2) |

**Table S17.** Results of nested cross-validation for the six features calculated on EEG filtered to alpha band (8-12.5 Hz), with no dimensionality reduction. For each of the ten repeats of fold selection, each individual is classified exactly once as part of one of the ten non-overlapping testing folds. Means and standard deviations are calculated for outcome measures across these ten repeats.

| | Full Sample | | | | Subset (min. 4 epochs) | | |
|---|---|---|---|---|---|---|---|
| **Model** | **Balanced Accuracy mean (SD) (%)** | **Sensitivity (Epilepsy) mean (SD) (%)** | **Sensitivity (FDS) mean (SD) (%)** | | **Balanced Accuracy mean (SD) (%)** | **Sensitivity (Epilepsy) mean (SD) (%)** | **Sensitivity (FDS) mean (SD) (%)** |
| Dummy Classifier | 51.7 (3.6) | 68.4 (5.2) | 34.9 (5.4) | | 52.1 (4.9) | 68.1 (4.7) | 36.2 (7.7) |
| Decision Tree | 47.3 (3.6) | 44.8 (4.1) | 49.7 (5.7) | | 50.6 (3.3) | 73.3 (6.6) | 27.8 (3.3) |
| Support Vector Machine (Sigmoid kernel) | 46.6 (2.4) | 50.9 (4.4) | 42.3 (4.2) | | 45.3 (5.0) | 57.5 (6.0) | 33.1 (7.5) |
| XGBoost | 46.5 (3.1) | 49.1 (6.0) | 44.0 (4.4) | | 40.9 (3.3) | 51.9 (5.6) | 29.8 (5.1) |
| Linear Discriminant Analysis | 46.2 (3.5) | 50.1 (5.2) | 42.2 (6.4) | | 46.4 (2.7) | 70.0 (3.5) | 22.9 (5.1) |
| RUSBoost | 46.0 (4.5) | 45.7 (5.3) | 46.3 (8.8) | | 41.1 (3.5) | 46.5 (7.6) | 35.8 (5.7) |
| Support Vector Machine (RBF kernel) | 45.7 (1.8) | 77.3 (3.0) | 14.1 (2.9) | | 44.4 (3.5) | 82.8 (4.1) | 6.0 (5.2) |
| Support Vector Machine (Linear kernel) | 45.5 (3.0) | 51.5 (5.2) | 39.6 (6.1) | | 46.1 (2.5) | 80.7 (3.0) | 11.6 (4.5) |
| k-Nearest Neighbours | 45.3 (2.3) | 38.9 (5.3) | 51.6 (3.4) | | 44.1 (5.0) | 57.7 (7.8) | 30.4 (5.4) |
| Logistic Regression | 45.1 (3.7) | 50.0 (6.5) | 40.1 (6.7) | | 45.5 (1.9) | 72.3 (3.4) | 18.7 (3.9) |
| AdaBoost | 44.5 (3.0) | 47.1 (4.8) | 41.9 (4.6) | | 44.3 (4.9) | 47.0 (7.2) | 41.6 (5.3) |
| Multi-layer Perceptron | 42.5 (3.1) | 44.5 (3.8) | 40.4 (4.9) | | 45.6 (4.8) | 55.8 (7.1) | 35.3 (8.2) |
| Random Forest | 42.3 (4.0) | 43.9 (4.4) | 40.7 (7.9) | | 41.1 (3.6) | 64.4 (8.0) | 17.8 (4.9) |
| Gaussian Naive Bayes | 41.1 (3.5) | 51.7 (6.4) | 30.5 (3.7) | | 40.5 (3.5) | 67.2 (6.7) | 13.8 (3.7) |

**Table S18.** Results of nested cross-validation for the six features calculated on EEG filtered to beta band (13-30 Hz), with no dimensionality reduction. For each of the ten repeats of fold selection, each individual is classified exactly once as part of one of the ten non-overlapping testing folds. Means and standard deviations are calculated for outcome measures across these ten repeats.

| | **Full Sample** | | | | **Subset (min. 4 epochs)** | | |
|---|---|---|---|---|---|---|---|
| **Model** | **Balanced Accuracy mean (SD) (%)** | **Sensitivity (Epilepsy) mean (SD) (%)** | **Sensitivity (FDS) mean (SD) (%)** | | **Balanced Accuracy mean (SD) (%)** | **Sensitivity (Epilepsy) mean (SD) (%)** | **Sensitivity (FDS) mean (SD) (%)** |
| Support Vector Machine (Sigmoid kernel) | 52.8 (3.8) | 56.5 (5.0) | 49.0 (6.3) | | 46.0 (4.2) | 58.9 (5.8) | 33.1 (6.0) |
| Dummy Classifier | 51.7 (3.6) | 68.4 (5.2) | 34.9 (5.4) | | 52.1 (4.9) | 68.1 (4.7) | 36.2 (7.7) |
| Gaussian Naive Bayes | 51.5 (2.8) | 64.8 (4.9) | 38.2 (5.6) | | 42.0 (3.1) | 67.2 (4.7) | 16.9 (4.4) |
| RUSBoost | 51.0 (3.0) | 50.0 (2.6) | 51.9 (4.5) | | 48.6 (5.2) | 55.4 (7.3) | 41.8 (5.8) |
| Linear Discriminant Analysis | 50.6 (3.1) | 53.5 (4.3) | 47.8 (3.3) | | 52.5 (5.9) | 70.9 (4.7) | 34.2 (10.8) |
| k-Nearest Neighbours | 50.3 (4.3) | 62.1 (6.0) | 38.5 (4.9) | | 50.0 (3.9) | 72.6 (6.9) | 27.3 (5.4) |
| Decision Tree | 50.1 (3.4) | 49.5 (4.3) | 50.8 (4.1) | | 49.1 (4.0) | 54.6 (4.9) | 43.6 (5.1) |
| Logistic Regression | 49.8 (2.7) | 53.7 (4.1) | 45.9 (3.8) | | 52.0 (5.5) | 72.8 (5.5) | 31.1 (9.9) |
| AdaBoost | 49.6 (2.9) | 47.6 (4.7) | 51.5 (4.1) | | 51.0 (3.1) | 61.1 (7.3) | 40.9 (6.3) |
| Support Vector Machine (RBF kernel) | 49.5 (3.7) | 56.4 (4.3) | 42.6 (5.4) | | 54.0 (5.5) | 66.5 (8.0) | 41.6 (6.8) |
| Support Vector Machine (Linear kernel) | 49.4 (1.1) | 57.2 (1.8) | 41.6 (2.5) | | 46.9 (3.0) | 78.2 (3.6) | 15.6 (7.0) |
| Multi-layer Perceptron | 47.9 (4.1) | 47.3 (5.4) | 48.5 (5.8) | | 49.0 (4.8) | 56.1 (5.7) | 41.8 (6.3) |
| XGBoost | 45.6 (3.8) | 46.0 (5.3) | 45.2 (3.6) | | 51.1 (3.7) | 61.1 (6.1) | 41.1 (4.8) |
| Random Forest | 45.3 (3.9) | 53.3 (7.1) | 37.3 (5.7) | | 46.7 (3.5) | 64.9 (5.8) | 28.4 (5.7) |

### Section 5. Features medians and statistical comparisons

**Table S19.** The median and interquartile range of the features (mean network measures for each person) for the epilepsy and FDS groups, and test statistic and p-value for a Mann-Whitney U-test comparing the distribution of each feature in the two diagnostic groups; for both the full sample and the subset of people with at least four epochs of EEG.

| | **Full sample** | | | | **Subset (min. 4 epochs)** | | | |
|---|---|---|---|---|---|---|---|---|
| **Measure** | **Epilepsy group median (IQR)** | **FDS group median (IQR)** | **Test Statistic U** | **p-value** | **Epilepsy group median (IQR)** | **FDS group median (IQR)** | **Test Statistic U** | **p-value** |
| Mean Clustering Coefficient | 0.0569 (0.00322) | 0.0572 (0.00425) | 2967 | 0.380 | 0.0568 (0.00336) | 0.0562 (0.00366) | 1287 | 0.978 |
| Mean Functional Connectivity | 0.376 (0.0449) | 0.366 (0.0573) | 2527 | 0.421 | 0.374 (0.0506) | 0.359 (0.048) | 1065 | 0.144 |
| Efficiency | 0.144 (0.0035) | 0.145 (0.00508) | 3096 | 0.170 | 0.144 (0.00263) | 0.146 (0.00386) | 1535 | 0.089 |
| Trophic Incoherence | 0.884 (0.0266) | 0.883 (0.0333) | 2545 | 0.461 | 0.885 (0.027) | 0.883 (0.0286) | 1136 | 0.325 |
| Critical Coupling | 2.2 (0.069) | 2.19 (0.0446) | 2520 | 0.405 | 2.2 (0.0758) | 2.19 (0.0379) | 1123 | 0.284 |
| Betweenness Centrality | 27.1 (1.87) | 27.7 (2.08) | 3129.5 | 0.133 | 27 (1.6) | 27.4 (1.85) | 1538 | 0.086 |

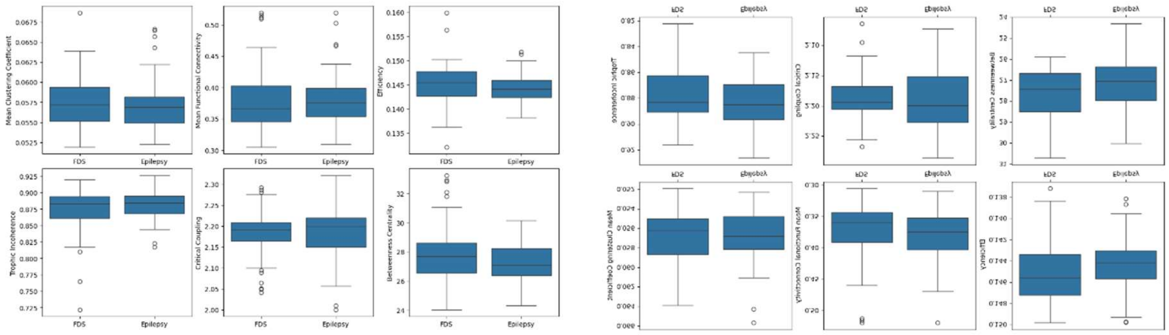

(a) Full sample

(b) Subset with at least four epochs

**Figure S1.** Box-plots showing the distributions (median, quartiles) of mean network measures (mean clustering coefficient, mean functional connectivity, efficiency, trophic incoherence, critical coupling, betweenness centrality) of individuals in (a) full sample and (b) subset with at least four epochs of EEG. Outliers (values more than 1.5 times the interquartile range from the first and third quartiles) are indicated with circles.

#### Section 6. Generalised Linear Model of potential confounders

**Table S20.** Generalised linear model results. Diagnostic classification is regressed against a binary variable indicating the presence of one or multiple comorbid diagnoses, sex as a binary variable, and age at the time of EEG recording as an integer number of years. Output shows the regression coefficient (β), alongside the 95% confidence bounds for the coefficient, standard error (SE), z-score (z), and p-value.

| **Confounder** | **β** | **95% CI (lower)** | **95% CI (upper)** | **SE** | **z** | **p-value** |
|---|---|---|---|---|---|---|
| Intercept | -0.1926 | -1.478 | 1.093 | 0.656 | -0.294 | 0.769 |
| Presence of comorbidities | 0.7373 | -0.128 | 1.603 | 0.442 | 1.669 | 0.095 |
| Sex | 0.2703 | -0.572 | 1.113 | 0.430 | 0.629 | 0.530 |
| Age | -0.0143 | -0.048 | 0.019 | 0.017 | -0.842 | 0.400 |
